## Supplementary material for "Preventing COVID-19 spread in closed facilities by regular testing of employees – an efficient intervention in long-term care facilities and prisons?": Mathematical Appendix

### Appendix: Mathematical description

For simplicity, we describe the model in terms of LTCFs. However, the model is applicable to closed facilities in general and the term LTCF can be replaced by e.g., Prison.

A population of $N$ is divided into three interacting sub-populations, (i) the immobile risk group, i.e., residents of LTCFs, (ii) the employees (staff) working in LTCFs, who are in close contact with the risk group, and (iii) the general population, i.e., the rest of the population. We use Ri, St, Ge as super and sub-scripts to refer to these sub-populations in the following. The sub-populations are not equally sized. The general population size is much larger than the risk group and the group of LTCF employees.

$$S^{(\mathrm{Ge})}(t), \quad S^{(\mathrm{St})}(t), \quad S^{(\mathrm{Ri})}(t), \tag{1}$$

are the numbers of susceptible individuals in the general, staff and risk group sub-populations at time $t$.

Susceptible individuals become infected by contacts with infectious individuals. Contacts across the sub-populations (Ge, St, Ri) are possible, however subject to the inherent interactions between these groups. Infected individuals progress from a latent ($E$), to a prodromal ($P$), to a fully ($I$), and finally to a late ($L$) phase of the disease, before the either recover ($R$) and become immune or die ($D$). The relative infectiousness in the prodromal, fully contagious and late infectious states are $c_P$, $c_I$, and $c_L$, respectively. The average duration an infected individual spends in the respective stages are denoted by $D_E$, $D_P$, $D_I$, and $D_L$. In simple SEIR models, implicitly, these durations are exponentially distributed, implying that the variance of the durations are given by $D_E^2$, $D_P^2$, $D_I^2$, $D_L^2$, respectively. To mitigate this naive dynamics, we partition each phase of the infection into consecutive identical sub-stages, that yield Erlang-distributed durations. Let $n_E$, $n_P$, $n_I$, and $n_L$ be the number of latent, prodromal, fully contagious and late infective sub-stages. The average time spent in each sub-stage of phase $H \in \{E, P, I, L\}$ of the disease is hence $D_H/n_H$. Therefore, the average duration spend in phase $H$ is $D_H$, however, the variance of the duration is $D_H^2/n_H$. Thus, the number of sub-stages, shrinks the variance of the respective durations. The sub-stages are denoted by $H_k^{(.)}$, for $k = 1, \ldots, n_H$, where (.) is a placeholder for the respective sub-populations.

The average duration spent in the latent, prodromal, fully contagious, and late infectious sub-stages are hence

$$\frac{D_E}{n_E} = \frac{1}{\varepsilon}, \quad \frac{D_P}{n_P} = \frac{1}{\varphi}, \quad \frac{D_I}{n_I} = \frac{1}{\gamma}, \quad \frac{D_L}{n_L} = \frac{1}{\delta}, \tag{2}$$

with $\varepsilon$, $\varphi$, $\gamma$, $\delta$ being the rates of change between compartments.

The LTCF employees will be tested for COVID-19 on a regular basis, and isolated if tested positive. More precisely, staff is tested at a rate $\xi$, i.e., each staff is tested $1/\xi$ times per time unit. We assume that the test is 100% specific, i.e., there are no false-positive test results, reflecting PCR- or CRISPR-based tests [?, ?]. Thus, testing does not need to be modelled explicitly in the susceptible staff population. The test, however, is not 100% sensitive, i.e., false-negative results occur. We will use $\mathrm{St}, +$ and $\mathrm{St}, -$ as super- and sub-scripts to refer to infected staff who are tested positive and not yet positive (either not tested or false-negative), respectively. The sensitivity of the test depends on the phase of the infection. Let $s_E$, $s_P$, $s_I$, $s_L$, denote the test's sensitivity in the latent, prodromal, fully contagious and late infective phases.

Importantly, test results are not obtained immediately, but with a time delay. The waiting time for test results is $1/\alpha$. The notation $\mathrm{St}, *$ is used as super- and sub-scripts to refer to staff awaiting the test results, whose test result will be positive.

The total number of latent infected individuals in the general population is

$$E_{\mathrm{Sum}}^{(\mathrm{Ge})}(t) = \sum_{k=1}^{n_E} E_k^{(\mathrm{Ge})}(t). \tag{3a}$$

In the population of LTCF staff, it is necessary to distinguish between those that have not yet been tested positive, those that will be tested positive but are still waiting the test results and those that have tested positive. The numbers of latently infected individuals in these sub-populations are

$$E_{\mathrm{Sum}}^{(\mathrm{St},-)}(t) = \sum_{k=1}^{n_E} E_k^{(\mathrm{St},-)}(t), \tag{3b}$$

$$E_{\mathrm{Sum}}^{(\mathrm{St},*)}(t) = \sum_{k=1}^{n_E} E_k^{(\mathrm{St},*)}(t), \tag{3c}$$

$$E_{\mathrm{Sum}}^{(\mathrm{St},+)}(t) = \sum_{k=1}^{n_E} E_k^{(\mathrm{St},+)}(t). \tag{3d}$$

The number of latent infections in the risk group is

$$E_{\mathrm{Sum}}^{(\mathrm{Ri})}(t) = \sum_{k=1}^{n_E} E_k^{(\mathrm{Ri})}(t). \tag{3e}$$

Similarly, the total numbers of prodromal infections in the sub-populations are

$$P_{\mathrm{Sum}}^{(\mathrm{Ge})}(t) = \sum_{k=1}^{n_P} P_k^{(\mathrm{Ge})}(t), \tag{4a}$$

$$P_{\mathrm{Sum}}^{(\mathrm{St},-)}(t) = \sum_{k=1}^{n_P} P_k^{(\mathrm{St},-)}(t), \tag{4b}$$

$$P_{\mathrm{Sum}}^{(\mathrm{St},*)}(t) = \sum_{k=1}^{n_P} P_k^{(\mathrm{St},*)}(t), \tag{4c}$$

$$P_{\mathrm{Sum}}^{(\mathrm{St},+)}(t) = \sum_{k=1}^{n_P} P_k^{(\mathrm{St},+)}(t), \tag{4d}$$

$$P_{\mathrm{Sum}}^{(\mathrm{Ri})}(t) = \sum_{k=1}^{n_P} P_k^{(\mathrm{Ri})}(t). \tag{4e}$$

The numbers of fully contagious individuals in the respective sub-populations are

$$I_{\mathrm{Sum}}^{(\mathrm{Ge})}(t) = \sum_{k=1}^{n_I} I_k^{(\mathrm{Ge})}(t), \tag{5a}$$

$$I_{\mathrm{Sum}}^{(\mathrm{St},-)}(t) = \sum_{k=1}^{n_I} I_k^{(\mathrm{St},-)}(t), \tag{5b}$$

$$I_{\mathrm{Sum}}^{(\mathrm{St},*)}(t) = \sum_{k=1}^{n_I} I_k^{(\mathrm{St},*)}(t), \tag{5c}$$

$$I_{\mathrm{Sum}}^{(\mathrm{St},+)}(t) = \sum_{k=1}^{n_I} I_k^{(\mathrm{St},+)}(t), \tag{5d}$$

$$I_{\mathrm{Sum}}^{(\mathrm{Ri})}(t) = \sum_{k=1}^{n_I} I_k^{(\mathrm{Ri})}(t), \tag{5e}$$

while those of the late infectious are

$$L_{\mathrm{Sum}}^{(\mathrm{Ge})}(t) = \sum_{k=1}^{n_L} L_k^{(\mathrm{Ge})}(t), \tag{6a}$$

$$L_{\mathrm{Sum}}^{(\mathrm{St},-)}(t) = \sum_{k=1}^{n_L} L_k^{(\mathrm{St},-)}(t), \tag{6b}$$

$$L_{\mathrm{Sum}}^{(\mathrm{St},*)}(t) = \sum_{k=1}^{n_L} L_k^{(\mathrm{St},*)}(t), \tag{6c}$$

$$L_{\mathrm{Sum}}^{(\mathrm{St},+)}(t) = \sum_{k=1}^{n_L} L_k^{(\mathrm{St},+)}(t), \tag{6d}$$

$$L_{\mathrm{Sum}}^{(\mathrm{Ri})}(t) = \sum_{k=1}^{n_L} L_k^{(\mathrm{Ri})}(t). \tag{6e}$$

COVID-19 testing does not need to be modelled among recovered staff, since they are immune. Note that also recovered individuals will be tested regularly, if their infection was undetected. To assess the amount of unnecessary performed tests, it is hence important to distinguish between the various groups also among recovered individuals. The total numbers of recovered individuals in the general population, the staff $(\mathrm{St},+;\ \mathrm{St},*;\ \mathrm{St},-)$ and the risk group are $R^{(\mathrm{St},+)}$, $R^{(\mathrm{St},*)}$, $R^{(\mathrm{St},-)}$, and $R^{(\mathrm{Ri})}$, while the number of deaths that occurred until time $t$ in the sub-populations are $D^{(\mathrm{St},-)}$, $D^{(\mathrm{St},*)}$, $D^{(\mathrm{St},-)}$, and $D^{(\mathrm{Ri})}$.

A fraction $f_{\mathrm{Sick}}$ of fully contagious individuals develop symptoms, i.e., the get sick. The fraction is higher in the risk group and denoted by ${f_{\mathrm{Sick}}}^{(\mathrm{Ri})}$. Fractions $f_{\mathrm{Dead}}$ in the general and LTCF staff sub-populations and $f_{\mathrm{Dead}}^{(\mathrm{Ri})}$ in the risk group of symptomatic individuals will ultimately die.

Thus the numbers of symptomatic infections in the fully contagious states in the various sub-populations are

$$I_{\mathrm{Sick}}^{(\mathrm{Ge})}(t) = f_{\mathrm{Sick}} I_{\mathrm{Sum}}^{(\mathrm{Ge})}(t), \tag{7a}$$

$$I_{\mathrm{Sick}}^{(\mathrm{St},-)}(t) = f_{\mathrm{Sick}} I_{\mathrm{Sum}}^{(\mathrm{St},-)}(t), \tag{7b}$$

$$I_{\mathrm{Sick}}^{(\mathrm{St},*)}(t) = f_{\mathrm{Sick}} I_{\mathrm{Sum}}^{(\mathrm{St},*)}(t), \tag{7c}$$

$$I_{\mathrm{Sick}}^{(\mathrm{St},+)}(t) = f_{\mathrm{Sick}} I_{\mathrm{Sum}}^{(\mathrm{St},+)}(t), \tag{7d}$$

$$I_{\mathrm{Sick}}^{(\mathrm{Ri})}(t) = f_{\mathrm{Sick}} I_{\mathrm{Sum}}^{(\mathrm{Ri})}(t), \tag{7e}$$

and the number of individuals in the late infectious states that are/were symptomatic are

$$L^{(\mathrm{Ge})}_{\mathrm{Sick}}(t) = f_{\mathrm{Sick}} L^{(\mathrm{Ge})}_{\mathrm{Sum}}(t), \tag{8a}$$

$$L^{(\mathrm{St},-)}_{\mathrm{Sick}}(t) = f_{\mathrm{Sick}} L^{(\mathrm{St},-)}_{\mathrm{Sum}}(t), \tag{8b}$$

$$L^{(\mathrm{St},*)}_{\mathrm{Sick}}(t) = f_{\mathrm{Sick}} L^{(\mathrm{St},*)}_{\mathrm{Sum}}(t), \tag{8c}$$

$$L^{(\mathrm{St},+)}_{\mathrm{Sick}}(t) = f_{\mathrm{Sick}} L^{(\mathrm{St},+)}_{\mathrm{Sum}}(t), \tag{8d}$$

$$L^{(\mathrm{Ri})}_{\mathrm{Sick}}(t) = f_{\mathrm{Sick}} L^{(\mathrm{Ri})}_{\mathrm{Sum}}(t). \tag{8e}$$

Case isolation mechanisms are sustained during the time interval from $t_{\mathrm{Iso}_1}$ to $t_{\mathrm{Iso}_2}$. In the risk group symptomatic infections will be isolated in quarantine, as LTCFs are equipped for these purposes. In the rest of the population a fraction $f_{\mathrm{Iso}}$ of symptomatic infections gets hospitalized and will be put into quarantine wards until their maximum capacity $Q_{\max}$ is reached. In the latter case, they will go into home isolation. Whereas quarantine wards prevent all infectious contacts, home isolation prevents only a fraction $p_{\mathrm{Home}}$ of contacts. Importantly, no individual in home isolation has any contact with the risk group, i.e., infective contacts between an individual in home isolation and the risk group cannot occur in the time interval from $t_{\mathrm{Iso}_1}$ to $t_{\mathrm{Iso}_2}$. Staff, positively tested for COVID-19, even if asymptomatic, also go into isolation in the quarantine wards or home isolation if the wards are occupied. Individuals stay in isolation until they recover or die, during the time interval from $t_{\mathrm{Iso}_1}$ to $t_{\mathrm{Iso}_2}$.

The number of individuals from the general and staff sub-populations in isolation facilities at time $t$ are

$$\begin{aligned} Q(t) = & f_{\mathrm{Iso}} \Big( I^{(\mathrm{Ge})}_{\mathrm{Sick}}(t) + L^{(\mathrm{Ge})}_{\mathrm{Sick}}(t) + I^{(\mathrm{St},-)}_{\mathrm{Sick}}(t) + L^{(\mathrm{St},-)}_{\mathrm{Sick}}(t) + I^{(\mathrm{St},*)}_{\mathrm{Sick}}(t) + L^{(\mathrm{St},*)}_{\mathrm{Sick}}(t) \Big) \\ & + E^{(\mathrm{St},+)}_{\mathrm{Sum}}(t) + P^{(\mathrm{St},+)}_{\mathrm{Sum}}(t) + I^{(\mathrm{St},+)}_{\mathrm{Sum}}(t) + L^{(\mathrm{St},+)}_{\mathrm{Sum}}(t). \end{aligned} \tag{9}$$

Thus the numbers of latently infected and prodromal staff in quarantine wards are

$$E^{(\mathrm{St},+)}_{\mathrm{Iso}}(t) = \begin{cases} E^{(\mathrm{St},+)}_{\mathrm{Sum}}(t) & \text{if } t_{\mathrm{Iso}_1} \le t \le t_{\mathrm{Iso}_2} \quad \text{and} \quad Q(t) \le Q_{\max}, \\ E^{(\mathrm{St},+)}_{\mathrm{Sum}}(t) \dfrac{Q_{\max}}{Q(t)} & \text{if } t_{\mathrm{Iso}_1} \le t \le t_{\mathrm{Iso}_2} \quad \text{and} \quad Q(t) > Q_{\max}, \\ 0 & \text{otherwise,} \end{cases} \tag{10}$$

and

$$P^{(\mathrm{St},+)}_{\mathrm{Iso}}(t) = \begin{cases} P^{(\mathrm{St},+)}_{\mathrm{Sum}}(t) & \text{if } t_{\mathrm{Iso}_1} \le t \le t_{\mathrm{Iso}_2} \quad \text{and} \quad Q(t) \le Q_{\max}, \\ P^{(\mathrm{St},+)}_{\mathrm{Sum}}(t) \dfrac{Q_{\max}}{Q(t)} & \text{if } t_{\mathrm{Iso}_1} \le t \le t_{\mathrm{Iso}_2} \quad \text{and} \quad Q(t) > Q_{\max}, \\ 0 & \text{otherwise,} \end{cases} \tag{11}$$

The numbers of fully contagious individuals in quarantine wards in the various sub-populations are

$$I_{\text{Iso}}^{(\text{Ge})}(t) = \begin{cases} f_{\text{Iso}} I_{\text{Sick}}^{(\text{Ge})}(t) & \text{if } t_{\text{Iso}_1} \le t \le t_{\text{Iso}_2} \quad \text{and} \quad Q(t) \le Q_{\max}, \\ f_{\text{Iso}} I_{\text{Sick}}^{(\text{Ge})}(t) \dfrac{Q_{\max}}{Q(t)} & \text{if } t_{\text{Iso}_1} \le t \le t_{\text{Iso}_2} \quad \text{and} \quad Q(t) > Q_{\max}, \\ 0 & \text{otherwise,} \end{cases} \tag{12a}$$

$$I_{\text{Iso}}^{(\text{St},-)}(t) = \begin{cases} f_{\text{Iso}} I_{\text{Sick}}^{(\text{St},-)}(t) & \text{if } t_{\text{Iso}_1} \le t \le t_{\text{Iso}_2} \quad \text{and} \quad Q(t) \le Q_{\max}, \\ f_{\text{Iso}} I_{\text{Sick}}^{(\text{St},-)}(t) \dfrac{Q_{\max}}{Q(t)} & \text{if } t_{\text{Iso}_1} \le t \le t_{\text{Iso}_2} \quad \text{and} \quad Q(t) > Q_{\max}, \\ 0 & \text{otherwise,} \end{cases} \tag{12b}$$

$$I_{\text{Iso}}^{(\text{St},*)}(t) = \begin{cases} f_{\text{Iso}} I_{\text{Sick}}^{(\text{St},*)}(t) & \text{if } t_{\text{Iso}_1} \le t \le t_{\text{Iso}_2} \quad \text{and} \quad Q(t) \le Q_{\max}, \\ f_{\text{Iso}} I_{\text{Sick}}^{(\text{St},*)}(t) \dfrac{Q_{\max}}{Q(t)} & \text{if } t_{\text{Iso}_1} \le t \le t_{\text{Iso}_2} \quad \text{and} \quad Q(t) > Q_{\max}, \\ 0 & \text{otherwise,} \end{cases} \tag{12c}$$

$$I_{\text{Iso}}^{(\text{St},+)}(t) = \begin{cases} I_{\text{Sum}}^{(\text{St},+)}(t) & \text{if } t_{\text{Iso}_1} \le t \le t_{\text{Iso}_2} \quad \text{and} \quad Q(t) \le Q_{\max}, \\ I_{\text{Sum}}^{(\text{St},+)}(t) \dfrac{Q_{\max}}{Q(t)} & \text{if } t_{\text{Iso}_1} \le t \le t_{\text{Iso}_2} \quad \text{and} \quad Q(t) > Q_{\max}, \\ 0 & \text{otherwise,} \end{cases} \tag{12d}$$

$$I_{\text{Iso}}^{(\text{Ri})}(t) = \begin{cases} I_{\text{Sick}}^{(\text{Ri})}(t) & \text{if } t_{\text{Iso}_1} \le t \le t_{\text{Iso}_2}, \\ 0 & \text{otherwise.} \end{cases} \tag{12e}$$

The numbers of individuals in the late infected states in quarantine wards are

$$L_{\text{Iso}}^{(\text{Ge})}(t) = \begin{cases} f_{\text{Iso}} L_{\text{Sick}}^{(\text{Ge})}(t) & \text{if } t_{\text{Iso}_1} \le t \le t_{\text{Iso}_2} \quad \text{and} \quad Q(t) \le Q_{\max}, \\ f_{\text{Iso}} L_{\text{Sick}}^{(\text{Ge})}(t) \dfrac{Q_{\max}}{Q(t)} & \text{if } t_{\text{Iso}_1} \le t \le t_{\text{Iso}_2} \quad \text{and} \quad Q(t) > Q_{\max}, \\ 0 & \text{otherwise,} \end{cases} \tag{13a}$$

$$L_{\text{Iso}}^{(\text{St},-)}(t) = \begin{cases} f_{\text{Iso}} L_{\text{Sick}}^{(\text{St},-)}(t) & \text{if } t_{\text{Iso}_1} \le t \le t_{\text{Iso}_2} \quad \text{and} \quad Q(t) \le Q_{\max}, \\ f_{\text{Iso}} L_{\text{Sick}}^{(\text{St},-)}(t) \dfrac{Q_{\max}}{Q(t)} & \text{if } t_{\text{Iso}_1} \le t \le t_{\text{Iso}_2} \quad \text{and} \quad Q(t) > Q_{\max}, \\ 0 & \text{otherwise,} \end{cases} \tag{13b}$$

$$L_{\text{Iso}}^{(\text{St},*)}(t) = \begin{cases} f_{\text{Iso}} L_{\text{Sick}}^{(\text{St},*)}(t) & \text{if } t_{\text{Iso}_1} \le t \le t_{\text{Iso}_2} \quad \text{and} \quad Q(t) \le Q_{\max}, \\ f_{\text{Iso}} L_{\text{Sick}}^{(\text{St},*)}(t) \dfrac{Q_{\max}}{Q(t)} & \text{if } t_{\text{Iso}_1} \le t \le t_{\text{Iso}_2} \quad \text{and} \quad Q(t) > Q_{\max}, \\ 0 & \text{otherwise,} \end{cases} \tag{13c}$$

$$L_{\text{Iso}}^{(\text{St},+)}(t) = \begin{cases} L_{\text{Sum}}^{(\text{St},+)}(t) & \text{if } t_{\text{Iso}_1} \le t \le t_{\text{Iso}_2} \quad \text{and} \quad Q(t) \le Q_{\max}, \\ L_{\text{Sum}}^{(\text{St},+)}(t) \dfrac{Q_{\max}}{Q(t)} & \text{if } t_{\text{Iso}_1} \le t \le t_{\text{Iso}_2} \quad \text{and} \quad Q(t) > Q_{\max}, \\ 0 & \text{otherwise,} \end{cases} \tag{13d}$$

$$L_{\text{Iso}}^{(\text{Ri})}(t) = \begin{cases} L_{\text{Sick}}^{(\text{Ri})}(t) & \text{if } t_{\text{Iso}_1} \le t \le t_{\text{Iso}_2}, \\ 0 & \text{otherwise.} \end{cases} \tag{13e}$$

Furthermore, the numbers of latently infected and prodromal positively tested staff in home isolation are given by

$$E^{(\mathrm{St},+)}_{\mathrm{Home}}(t) = \begin{cases} \left(1 - \dfrac{Q_{\max}}{Q(t)}\right) E^{(\mathrm{St},+)}_{\mathrm{Sum}}(t) & \text{if } t_{\mathrm{Iso}_1} \le t \le t_{\mathrm{Iso}_2} \text{ and } Q(t) > Q_{\max}, \\ 0 & \text{otherwise,} \end{cases} \tag{14}$$

$$P^{(\mathrm{St},+)}_{\mathrm{Home}}(t) = \begin{cases} \left(1 - \dfrac{Q_{\max}}{Q(t)}\right) P^{(\mathrm{St},+)}_{\mathrm{Sum}}(t) & \text{if } t_{\mathrm{Iso}_1} \le t \le t_{\mathrm{Iso}_2} \text{ and } Q(t) > Q_{\max}, \\ 0 & \text{otherwise,} \end{cases} \tag{15}$$

The total numbers of fully contagious individuals in home isolation at time $t$ is

$$I^{(\mathrm{Ge})}_{\mathrm{Home}}(t) = \begin{cases} \left(1 - \dfrac{Q_{\max}}{Q(t)}\right) f_{\mathrm{Iso}} I^{(\mathrm{Ge})}_{\mathrm{Sick}}(t) & \text{if } t_{\mathrm{Iso}_1} \le t \le t_{\mathrm{Iso}_2} \text{ and } Q(t) > Q_{\max}, \\ 0 & \text{otherwise,} \end{cases} \tag{16a}$$

$$I^{(\mathrm{St},-)}_{\mathrm{Home}}(t) = \begin{cases} \left(1 - \dfrac{Q_{\max}}{Q(t)}\right) f_{\mathrm{Iso}} I^{(\mathrm{St},-)}_{\mathrm{Sick}}(t) & \text{if } t_{\mathrm{Iso}_1} \le t \le t_{\mathrm{Iso}_2} \text{ and } Q(t) > Q_{\max}, \\ 0 & \text{otherwise,} \end{cases} \tag{16b}$$

$$I^{(\mathrm{St},*)}_{\mathrm{Home}}(t) = \begin{cases} \left(1 - \dfrac{Q_{\max}}{Q(t)}\right) f_{\mathrm{Iso}} I^{(\mathrm{St},*)}_{\mathrm{Sick}}(t) & \text{if } t_{\mathrm{Iso}_1} \le t \le t_{\mathrm{Iso}_2} \text{ and } Q(t) > Q_{\max}, \\ 0 & \text{otherwise,} \end{cases} \tag{16c}$$

$$I^{(\mathrm{St},+)}_{\mathrm{Home}}(t) = \begin{cases} \left(1 - \dfrac{Q_{\max}}{Q(t)}\right) I^{(\mathrm{St},+)}_{\mathrm{Sum}}(t) & \text{if } t_{\mathrm{Iso}_1} \le t \le t_{\mathrm{Iso}_2} \text{ and } Q(t) > Q_{\max}, \\ 0 & \text{otherwise,} \end{cases} \tag{16d}$$

and the total numbers of late infectious individuals in the sub-populations in home isolation are

$$L^{(\mathrm{Ge})}_{\mathrm{Home}}(t) = \begin{cases} \left(1 - \dfrac{Q_{\max}}{Q(t)}\right) f_{\mathrm{Iso}} L^{(\mathrm{Ge})}_{\mathrm{Sick}}(t) & \text{if } t_{\mathrm{Iso}_1} \le t \le t_{\mathrm{Iso}_2} \text{ and } Q(t) > Q_{\max}, \\ 0 & \text{otherwise,} \end{cases} \tag{17a}$$

$$L^{(\mathrm{St},-)}_{\mathrm{Home}}(t) = \begin{cases} \left(1 - \dfrac{Q_{\max}}{Q(t)}\right) f_{\mathrm{Iso}} I^{(\mathrm{St},-)}_{\mathrm{Sick}}(t) & \text{if } t_{\mathrm{Iso}_1} \le t \le t_{\mathrm{Iso}_2} \text{ and } Q(t) > Q_{\max}, \\ 0 & \text{otherwise,} \end{cases} \tag{17b}$$

$$L^{(\mathrm{St},*)}_{\mathrm{Home}}(t) = \begin{cases} \left(1 - \dfrac{Q_{\max}}{Q(t)}\right) f_{\mathrm{Iso}} L^{(\mathrm{St},*)}_{\mathrm{Sick}}(t) & \text{if } t_{\mathrm{Iso}_1} \le t \le t_{\mathrm{Iso}_2} \text{ and } Q(t) > Q_{\max}, \\ 0 & \text{otherwise,} \end{cases} \tag{17c}$$

$$L^{(\mathrm{St},+)}_{\mathrm{Home}}(t) = \begin{cases} \left(1 - \dfrac{Q_{\max}}{Q(t)}\right) L^{(\mathrm{St},+)}_{\mathrm{Sum}}(t) & \text{if } t_{\mathrm{Iso}_1} \le t \le t_{\mathrm{Iso}_2} \text{ and } Q(t) > Q_{\max}, \\ 0 & \text{otherwise.} \end{cases} \tag{17d}$$

Prodromal individuals in the general population and the risk group participate in infection. However, during the time case isolation measures are sustained, members of the general population are only allowed to enter the LTCF and get in contact with the risk group if tested negative. Assume a fraction $s_P^{(\mathrm{Ge})}$ of prodromal individuals in the general population cannot enter the LTCF because they obtained a negative test result (we neglect moving them into quarantine, because their number will be limited).

Hence, the number of prodromal individuals in the general population that can have infective contacts with the risk group is

$$P^{(\mathrm{Ge})}_{\mathrm{Eff,Ri}}(t) = P^{(\mathrm{Ge})}_{\mathrm{Sum}}(t)(1 - s_P^{(\mathrm{Ge})}). \tag{18a}$$

Prodromal staff also participate to infection, except those that are in isolation. The number of prodromal staff that can infect the general population and staff is

$$P_{\text{Eff}}^{(\text{St})}(t) = P_{\text{Sum}}^{(\text{St},-)}(t) + P_{\text{Sum}}^{(\text{St},*)}(t) + P_{\text{Sum}}^{(\text{St},+)}(t) - P_{\text{Iso}}^{(\text{St},+)}(t) - p_{\text{Home}} P_{\text{Home}}^{(\text{St},+)}(t). \quad (18b)$$

Positively-tested staff is isolated from the risk group, hence positively-tested prodromal staff cannot infect the risk group. Thus the number of prodromal staff that can infect the risk group is

$$P_{\text{Eff, Ri}}^{(\text{St})}(t) = P_{\text{Sum}}^{(\text{St},-)}(t) + P_{\text{Sum}}^{(\text{St},*)}(t). \quad (18c)$$

The number of fully contagious individuals in the general population that can infect susceptibles in the general and staff sub-populations is

$$I_{\text{Eff}}^{(\text{Ge})}(t) = I_{\text{Sum}}^{(\text{Ge})}(t) - I_{\text{Iso}}^{(\text{Ge})}(t) - p_{\text{Home}} I_{\text{Home}}^{(\text{Ge})}(t). \quad (19a)$$

Infected individuals in home isolation are isolated from the risk group. The remaining individuals need to provide a negative test before entry into the LTCF. Let $s_I^{(\text{Ge})}$ be the fraction of fully contagious individuals in the general population not in isolation that want to enter the LTCF but obtained a positive test results. Again it is ignored that these individuals are put into isolation. Hence, the number of fully contagious individuals in the general population infecting the risk group is

$$I_{\text{Eff, Ri}}^{(\text{Ge})}(t) = (1 - s_I^{(\text{Ge})})\big(I_{\text{Sum}}^{(\text{Ge})}(t) - I_{\text{Iso}}^{(\text{Ge})}(t) - I_{\text{Home}}^{(\text{Ge})}(t)\big). \quad (19b)$$

Similarly, the number of fully contagious individuals in the staff sub-populations, participating to infectious contacts with the general population and staff, is

$$\begin{aligned} I_{\text{Eff}}^{(\text{St})}(t) =& I_{\text{Sum}}^{(\text{St},-)}(t) - I_{\text{Iso}}^{(\text{St},-)}(t) - p_{\text{Home}} I_{\text{Home}}^{(\text{St},-)}(t) \\ &+ I_{\text{Sum}}^{(\text{St},*)}(t) - I_{\text{Iso}}^{(\text{St},*)}(t) - p_{\text{Home}} I_{\text{Home}}^{(\text{St},*)}(t) \\ &+ I_{\text{Sum}}^{(\text{St},+)}(t) - I_{\text{Iso}}^{(\text{St},+)}(t) - p_{\text{Home}} I_{\text{Home}}^{(\text{St},+)}(t). \end{aligned} \quad (19c)$$

Any staff in isolation (in wards or at home) are isolated from the risk group. Hence, the numbers of fully contagious staff that can infect the risk group are

$$I_{\text{Eff, Ri}}^{(\text{St})}(t) = I_{\text{Sum}}^{(\text{St},-)}(t) - I_{\text{Iso}}^{(\text{St},-)}(t) - I_{\text{Home}}^{(\text{St},-)}(t) + I_{\text{Sum}}^{(\text{St},*)}(t) - I_{\text{Iso}}^{(\text{St},*)}(t) - I_{\text{Home}}^{(\text{St},*)}(t). \quad (19d)$$

The number of fully contagious individuals in the risk group participating in infection is

$$I_{\text{Eff}}^{(\text{Ri})}(t) = I_{\text{Sum}}^{(\text{Ri})}(t) - I_{\text{Iso}}^{(\text{Ri})}(t). \quad (19e)$$

The number of late infected individuals in the general population participating in infective contacts with the general and staff populations is

$$L_{\text{Eff}}^{(\text{Ge})}(t) = L_{\text{Sum}}^{(\text{Ge})}(t) - L_{\text{Iso}}^{(\text{Ge})}(t) - p_{\text{Home}} L_{\text{Home}}^{(\text{Ge})}(t), \quad (20a)$$

whereas the number of late infectious individuals potentially infecting the risk group is

$$L_{\text{Eff, Ri}}^{(\text{Ge})}(t) = (1 - s_L^{(\text{Ge})})\big(L_{\text{Sum}}^{(\text{Ge})}(t) - L_{\text{Iso}}^{(\text{Ge})}(t) - L_{\text{Home}}^{(\text{Ge})}(t)\big), \quad (20b)$$

where $s_L^{(\mathrm{Ge})}$ is the probability that a late infected individual not in isolation that wants to enter the LTCF obtains a positive test result and is thus denied access. Again it is ignored to put these individuals into quarantine. The late infected staff population potentially infecting the general and staff populations is

$$\begin{aligned} L_{\mathrm{Eff}}^{(\mathrm{St})}(t) =& L_{\mathrm{Sum}}^{(\mathrm{St},-)}(t) - L_{\mathrm{Iso}}^{(\mathrm{St},-)}(t) - p_{\mathrm{Home}} L_{\mathrm{Home}}^{(\mathrm{St},-)}(t), \\ &+ L_{\mathrm{Sum}}^{(\mathrm{St},*)}(t) - L_{\mathrm{Iso}}^{(\mathrm{St},*)}(t) - p_{\mathrm{Home}} L_{\mathrm{Home}}^{(\mathrm{St},*)}(t), \\ &+ L_{\mathrm{Sum}}^{(\mathrm{St},+)}(t) - L_{\mathrm{Iso}}^{(\mathrm{St},+)}(t) - p_{\mathrm{Home}} L_{\mathrm{Home}}^{(\mathrm{St},+)}(t), \end{aligned} \tag{20c}$$

while those late infected staff potentially infecting the risk group amount to

$$\begin{aligned} L_{\mathrm{Eff,Ri}}^{(\mathrm{St})}(t) =& L_{\mathrm{Sum}}^{(\mathrm{St},-)}(t) - L_{\mathrm{Iso}}^{(\mathrm{St},-)}(t) - L_{\mathrm{Home}}^{(\mathrm{St},-)}(t) + L_{\mathrm{Sum}}^{(\mathrm{St},*)}(t) \\ &- L_{\mathrm{Iso}}^{(\mathrm{St},*)}(t) - L_{\mathrm{Home}}^{(\mathrm{St},*)}(t), \end{aligned} \tag{20d}$$

whereas the number of late infected in the risk group participating in infection is

$$L_{\mathrm{Eff}}^{(\mathrm{Ri})}(t) = L_{\mathrm{Sum}}^{(\mathrm{Ri})}(t) - L_{\mathrm{Iso}}^{(\mathrm{Ri})}(t). \tag{20e}$$

The basic reproduction number $R_0$ is the average number of infections caused by an average infected individual in a completely susceptible population, in which no interventions occur (cf. [?]). $R_0$ is assumed to fluctuate seasonally with a peak at time $t_{R_{0\max}}$, i.e.,

$$R_0(t) := \bar{R}_0 \left(1 + a \cos\left(2\pi \frac{t - t_{R_{0\max}}}{365}\right)\right), \tag{21}$$

where $\bar{R}_0$ is the seasonal average basic reproduction number, and $a$ $(0 \leq a \leq 1)$ determines the amplitude of seasonal fluctuations.

In populations, sub-divided into heterogeneous sub-populations, $\bar{R}_0$ is determined by the contact behaviors via the next generation matrix (see [?]). Contacts between sub-populations are not random, but reflect the specific interactions between them. Random encounters between the sub-populations are mediated by the symmetric mixing matrix

$$X(t) = \begin{pmatrix} x_{\mathrm{Ge,Ge}}(t) & x_{\mathrm{Ge,St}}(t) & x_{\mathrm{Ge,Ri}}(t) \\ x_{\mathrm{St,Ge}}(t) & x_{\mathrm{St,St}}(t) & x_{\mathrm{St,Ri}}(t) \\ x_{\mathrm{Ri,Ge}}(t) & x_{\mathrm{Ri,St}}(t) & x_{\mathrm{Ri,Ri}}(t) \end{pmatrix}, \tag{22}$$

whose entries are defined by (40), which corrects for the true contact behavior, described in detail below (see below section “Mathematical description of the demographic mixing matrix”). This matrix is time dependent because the contact behavior is further mediated by general contact reducing measure, e.g., curfews being sustained in certain time intervals. These contact-reducing measures depend on the characteristic of the sub-populations and are described below in detail.

The rates of infective contacts are determined by multiplying the mixing matrix with a rate, independent of the sub-populations. In the following these are referred to as contact rates. The contact rates for the prodromal, fully contagious and late infectious states are determined by the relative contagiousness, $c_P$, $c_I$, $c_L$, in these states and –

following [?] – are given by

$$\beta_P(t) := \frac{c_P \bar{R}_0^{(\mathrm{adj})}}{c_P D_P + c_I D_I + c_L D_L} \left(1 + a\cos\left(2\pi \frac{t - t_{R_{0_{\max}}}}{365}\right)\right), \tag{23a}$$

$$\beta_I(t) := \frac{c_I \bar{R}_0^{(\mathrm{adj})}}{c_P D_P + c_I D_I + c_L D_L} \left(1 + a\cos\left(2\pi \frac{t - t_{R_{0_{\max}}}}{365}\right)\right), \tag{23b}$$

$$\beta_L(t) := \frac{c_L \bar{R}_0^{(\mathrm{adj})}}{c_P D_P + c_I D_I + c_L D_L} \left(1 + a\cos\left(2\pi \frac{t - t_{R_{0_{\max}}}}{365}\right)\right), \tag{23c}$$

where $\bar{R}_0^{(\mathrm{adj})}$ is the adjusted average reproduction number that accounts for the contact behavior in the population.

Hence, the (internal) force of infection for susceptible in the general population becomes

$$\begin{aligned}\lambda_{\mathrm{Ge}}(t) =& \beta_P(t)\Big(x_{\mathrm{Ge,Ge}}(t) P_{\mathrm{Sum}}^{(\mathrm{Ge})}(t) + x_{\mathrm{Ge,St}}(t) P_{\mathrm{Eff}}^{(\mathrm{St})}(t) + x_{\mathrm{Ge,Ri}}(t) P_{\mathrm{Sum}}^{(\mathrm{Ri})}(t)\Big) \\ &+ \beta_I(t)\Big(x_{\mathrm{Ge,Ge}}(t) I_{\mathrm{Eff}}^{(\mathrm{Ge})}(t) + x_{\mathrm{Ge,St}}(t) I_{\mathrm{Eff}}^{(\mathrm{St})}(t) + x_{\mathrm{Ge,Ri}}(t) I_{\mathrm{Eff}}^{(\mathrm{Ri})}(t)\Big) \\ &+ \beta_L(t)\Big(x_{\mathrm{Ge,Ge}}(t) L_{\mathrm{Eff}}^{(\mathrm{Ge})}(t) + x_{\mathrm{Ge,St}}(t) L_{\mathrm{Eff}}^{(\mathrm{St})}(t) + x_{\mathrm{Ge,Ri}}(t) L_{\mathrm{Eff}}^{(\mathrm{Ri})}(t)\Big).\end{aligned} \tag{24}$$

The (internal) force of infection in the sub-populations of LTCF employees and the risk group are

$$\begin{aligned}\lambda_{\mathrm{St}}(t) =& \beta_P(t)\Big(x_{\mathrm{St,Ge}}(t) P_{\mathrm{Sum}}^{(\mathrm{Ge})}(t) + x_{\mathrm{St,St}}(t) P_{\mathrm{Eff}}^{(\mathrm{St})}(t) + x_{\mathrm{St,Ri}}(t) P_{\mathrm{Sum}}^{(\mathrm{Ri})}(t)\Big) \\ &+ \beta_I(t)\Big(x_{\mathrm{St,Ge}}(t) I_{\mathrm{Eff}}^{(\mathrm{Ge})}(t) + x_{\mathrm{St,St}}(t) I_{\mathrm{Eff}}^{(\mathrm{St})}(t) + x_{\mathrm{St,Ri}}(t) I_{\mathrm{Eff}}^{(\mathrm{Ri})}(t)\Big) \\ &+ \beta_L(t)\Big(x_{\mathrm{St,Ge}}(t) L_{\mathrm{Eff}}^{(\mathrm{Ge})}(t) + x_{\mathrm{St,St}}(t) L_{\mathrm{Eff}}^{(\mathrm{St})}(t) + x_{\mathrm{St,Ri}}(t) L_{\mathrm{Eff}}^{(\mathrm{Ri})}(t)\Big),\end{aligned} \tag{25}$$

and

$$\begin{aligned}\lambda_{\mathrm{Ri}}(t) =& x_{\mathrm{Ri,Ge}}(t)\Big(\beta_P(t) P_{\mathrm{Eff,Ri}}^{(\mathrm{Ge})}(t) + \beta_I(t) I_{\mathrm{Eff,Ri}}^{(\mathrm{Ge})}(t) + \beta_L(t) L_{\mathrm{Eff,Ri}}^{(\mathrm{Ge})}(t)\Big) \\ &+ \beta_P(t)\Big(x_{\mathrm{Ri,St}}(t) P_{\mathrm{Eff,Ri}}^{(\mathrm{St})}(t) + x_{\mathrm{Ri,Ri}}(t) P_{\mathrm{Sum}}^{(\mathrm{Ri})}(t)\Big) \\ &+ \beta_I(t)\Big(x_{\mathrm{Ri,St}}(t) I_{\mathrm{Eff,Ri}}^{(\mathrm{St})}(t) + x_{\mathrm{Ri,Ri}}(t) I_{\mathrm{Eff}}^{(\mathrm{Ri})}(t)\Big) \\ &+ \beta_L(t)\Big(x_{\mathrm{Ri,St}}(t) L_{\mathrm{Eff,Ri}}^{(\mathrm{St})}(t) + x_{\mathrm{Ri,Ri}}(t) L_{\mathrm{Eff}}^{(\mathrm{Ri})}(t)\Big),\end{aligned} \tag{26}$$

respectively.

Furthermore, we assume an external force of infection $\lambda_{\mathrm{Ext}}$ for the general population and LTCF staff. These are caused by infectious contacts with individuals from outside the population. We assume that the risk group has no contacts with these individuals.

Putting all together, the dynamics for the susceptible individuals become

$$\frac{dS^{(\mathrm{Ge})}}{dt} = -\Big(\lambda_{\mathrm{Ge}}(t) + \lambda_{\mathrm{Ext}}\Big)\frac{S^{(\mathrm{Ge})}(t)}{N}, \tag{27a}$$

$$\frac{dS^{(\mathrm{St})}}{dt} = -\Big(\lambda_{\mathrm{St}}(t) + \lambda_{\mathrm{Ext}}\Big)\frac{S^{(\mathrm{St})}}{N}, \tag{27b}$$

$$\frac{dS^{(\mathrm{Ri})}}{dt} = -\lambda_{\mathrm{Ri}}(t)\frac{S^{(\mathrm{Ri})}}{N}. \tag{27c}$$

The dynamics of the latently infected individuals in the respective sub-populations and Erlang states become

$$\frac{dE_1^{(\text{Ge})}}{dt} = \big(\lambda_{\text{Ge}}(t) + \lambda_{\text{Ext}}\big)\frac{S^{(\text{Ge})}(t)}{N} - \varepsilon E_1^{(\text{Ge})}(t), \tag{28a}$$

$$\frac{dE_k^{(\text{Ge})}}{dt} = \varepsilon E_{k-1}^{(\text{Ge})}(t) - \varepsilon E_k^{(\text{Ge})}(t) \qquad \text{for } 2 \leq k \leq n_E, \tag{28b}$$

$$\frac{dE_1^{(\text{St},-)}}{dt} = \big(\lambda_{\text{St}}(t)+\lambda_{\text{Ext}}\big)\frac{S^{(\text{St})}(t)}{N} - \varepsilon E_1^{(\text{St},-)}(t) - \xi s_E E_1^{(\text{St},-)}(t), \tag{28c}$$

$$\frac{dE_1^{(\text{St},*)}}{dt} = \xi s_E E_1^{(\text{St},-)}(t) - \varepsilon E_1^{(\text{St},*)}(t) - \alpha E_1^{(\text{St},*)}(t), \tag{28d}$$

$$\frac{dE_1^{(\text{St},+)}}{dt} = \alpha E_1^{(\text{St},*)}(t) - \varepsilon E_1^{(\text{St},+)}(t), \tag{28e}$$

$$\frac{dE_k^{(\text{St},-)}}{dt} = \varepsilon E_{k-1}^{(\text{St},-)}(t) - \varepsilon E_k^{(\text{St},-)}(t) - \xi s_E E_k^{(\text{St},-)}(t) \qquad \text{for } 2 \leq k \leq n_E, \tag{28f}$$

$$\frac{dE_k^{(\text{St},*)}}{dt} = \xi s_E E_k^{(\text{St},-)}(t) + \varepsilon E_{k-1}^{(\text{St},*)}(t) - \varepsilon E_k^{(\text{St},*)}(t) - \alpha E_k^{(\text{St},*)}(t) \qquad \text{for } 2 \leq k \leq n_E, \tag{28g}$$

$$\frac{dE_k^{(\text{St},+)}}{dt} = \alpha E_k^{(\text{St},*)}(t) + \varepsilon E_{k-1}^{(\text{St},+)}(t) - \varepsilon E_k^{(\text{St},+)}(t) \qquad \text{for } 2 \leq k \leq n_E, \tag{28h}$$

$$\frac{dE_1^{(\text{Ri})}}{dt} = \lambda_{\text{Ri}}(t)\frac{S^{(\text{Ri})}(t)}{N} - \varepsilon E_1^{(\text{Ri})}(t), \tag{28i}$$

$$\frac{dE_k^{(\text{Ri})}}{dt} = \varepsilon E_{k-1}^{(\text{Ri})}(t) - \varepsilon E_k^{(\text{Ri})}(t) \qquad \text{for } 2 \leq k \leq n_E. \tag{28j}$$

For the prodromal individuals the set of differential equations becomes

$$\frac{dP_1^{(\text{Ge})}}{dt} = \varepsilon E_{n_E}^{(\text{Ge})}(t) - \varphi P_1^{(\text{Ge})}(t), \tag{29a}$$

$$\frac{dP_k^{(\text{Ge})}}{dt} = \varphi P_{k-1}^{(\text{Ge})}(t) - \varphi P_k^{(\text{Ge})}(t) \qquad \text{for } 2 \leq k \leq n_P, \tag{29b}$$

$$\frac{dP_1^{(\text{St},-)}}{dt} = \varepsilon E_{n_E}^{(\text{St},-)}(t) - \varphi P_1^{(\text{St},-)}(t) - \xi s_P P_1^{(\text{St},-)}(t), \tag{29c}$$

$$\frac{dP_1^{(\text{St},*)}}{dt} = \varepsilon E_{n_E}^{(\text{St},*)}(t) + \xi s_P P_1^{(\text{St},-)}(t) - \varphi P_1^{(\text{St},*)}(t) - \alpha P_1^{(\text{St},*)}(t), \tag{29d}$$

$$\frac{dP_1^{(\text{St},+)}}{dt} = \varepsilon E_{n_E}^{(\text{St},+)}(t) + \alpha P_1^{(\text{St},*)}(t) - \varphi P_1^{(\text{St},+)}(t), \tag{29e}$$

$$\frac{dP_k^{(\text{St},-)}}{dt} = \varphi P_{k-1}^{(\text{St},-)}(t) - \varphi P_k^{(\text{St},-)}(t) - \xi s_P P_k^{(\text{St},-)}(t) \qquad \text{for } 2 \leq k \leq n_P, \tag{29f}$$

$$\frac{dP_k^{(\text{St},*)}}{dt} = \xi s_P P_k^{(\text{St},-)}(t) + \varphi P_{k-1}^{(\text{St},*)}(t) - \varphi P_k^{(\text{St},*)}(t) - \alpha P_k^{(\text{St},*)}(t) \qquad \text{for } 2 \leq k \leq n_P, \tag{29g}$$

$$\frac{dP_k^{(\text{St},+)}}{dt} = \alpha P_k^{(\text{St},*)}(t) + \varphi P_{k-1}^{(\text{St},+)}(t) - \varphi P_k^{(\text{St},+)}(t) \qquad \text{for } 2 \leq k \leq n_P, \tag{29h}$$

$$\frac{dP_1^{(\text{Ri})}}{dt} = \varepsilon E_{n_E}^{(\text{Ri})}(t) - \varphi P_1^{(\text{Ri})}(t), \tag{29i}$$

$$\frac{dP_k^{(\text{Ri})}}{dt} = \varphi P_{k-1}^{(\text{Ri})}(t) - \varphi P_k^{(\text{Ri})}(t) \qquad \text{for } \leq k \leq n_P. \tag{29j}$$

The dynamic of the fully infected population is described by the following differential equations

$$\frac{dI_1^{(\mathrm{Ge})}}{dt} = \varphi P_{n_P}^{(\mathrm{Ge})}(t) - \gamma I_1^{(\mathrm{Ge})}(t), \tag{30a}$$

$$\frac{dI_k^{(\mathrm{Ge})}}{dt} = \gamma I_{k-1}^{(\mathrm{Ge})}(t) - \gamma I_k^{(\mathrm{Ge})}(t) \qquad \text{for } 2 \le k \le n_I, \tag{30b}$$

$$\frac{dI_1^{(\mathrm{St},-)}}{dt} = \varphi P_{n_P}^{(\mathrm{St},-)}(t) - \gamma I_1^{(\mathrm{St},-)}(t) - \xi s_I I_1^{(\mathrm{St},-)}(t), \tag{30c}$$

$$\frac{dI_1^{(\mathrm{St},*)}}{dt} = \varphi P_{n_P}^{(\mathrm{St},*)}(t) + \xi s_I I_1^{(\mathrm{St},-)}(t) - \gamma I_1^{(\mathrm{St},*)}(t) - \alpha I_1^{(\mathrm{St},*)}(t), \tag{30d}$$

$$\frac{dI_1^{(\mathrm{St},+)}}{dt} = \varphi P_{n_P}^{(\mathrm{St},+)}(t) + \alpha I_1^{(\mathrm{St},*)}(t) - \gamma I_1^{(\mathrm{St},+)}(t), \tag{30e}$$

$$\frac{dI_k^{(\mathrm{St},-)}}{dt} = \gamma I_{k-1}^{(\mathrm{St},-)}(t) - \gamma I_k^{(\mathrm{St},-)}(t) - \xi s_I I_k^{(\mathrm{St},-)}(t) \qquad \text{for } 2 \le k \le n_I, \tag{30f}$$

$$\frac{dI_k^{(\mathrm{St},*)}}{dt} = \xi s_I I_k^{(\mathrm{St},-)}(t) + \gamma I_{k-1}^{(\mathrm{St},*)}(t) - \gamma I_k^{(\mathrm{St},*)}(t) - \alpha I_k^{(\mathrm{St},*)}(t) \qquad \text{for } 2 \le k \le n_I, \tag{30g}$$

$$\frac{dI_k^{(\mathrm{St},+)}}{dt} = \alpha I_k^{(\mathrm{St},*)}(t) + \gamma I_{k-1}^{(\mathrm{St},+)}(t) - \gamma I_k^{(\mathrm{St},+)}(t) \qquad \text{for } 2 \le k \le n_I, \tag{30h}$$

$$\frac{dI_1^{(\mathrm{Ri})}}{dt} = \varphi P_{n_P}^{(\mathrm{Ri})}(t) - \gamma I_1^{(\mathrm{Ri})}(t), \tag{30i}$$

$$\frac{dI_k^{(\mathrm{Ri})}}{dt} = \gamma I_{k-1}^{(\mathrm{Ri})}(t) - \gamma I_k^{(\mathrm{Ri})}(t) \qquad \text{for } 2 \le k \le n_I. \tag{30j}$$

The following differential equations describe the dynamic of the late infectious individuals

$$\frac{dL_1^{(\mathrm{Ge})}}{dt} = \gamma I_{n_I}^{(\mathrm{Ge})}(t) - \delta L_1^{(\mathrm{Ge})}(t), \tag{31a}$$

$$\frac{dL_k^{(\mathrm{Ge})}}{dt} = \delta L_{k-1}^{(\mathrm{Ge})}(t) - \delta L_k^{(\mathrm{Ge})}(t) \qquad \text{for } 2 \le k \le n_L, \tag{31b}$$

$$\frac{dL_1^{(\mathrm{St},-)}}{dt} = \gamma I_{n_I}^{(\mathrm{St},-)}(t) - \delta L_1^{(\mathrm{St},-)}(t) - \xi s_L L_1^{(\mathrm{St},-)}(t), \tag{31c}$$

$$\frac{dL_1^{(\mathrm{St},*)}}{dt} = \gamma I_{n_I}^{(\mathrm{St},*)}(t) + \xi s_L L_1^{(\mathrm{St},-)}(t) - \delta L_1^{(\mathrm{St},*)}(t) - \alpha L_1^{(\mathrm{St},*)}(t), \tag{31d}$$

$$\frac{dL_1^{(\mathrm{St},+)}}{dt} = \gamma I_{n_I}^{(\mathrm{St},+)}(t) + \alpha L_1^{(\mathrm{St},*)}(t) - \delta L_1^{(\mathrm{St},+)}(t), \tag{31e}$$

$$\frac{dL_k^{(\mathrm{St},-)}}{dt} = \delta L_{k-1}^{(\mathrm{St},-)}(t) - \delta L_k^{(\mathrm{St},-)}(t) - \xi s_L L_k^{(\mathrm{St},-)}(t) \qquad \text{for } 2 \le k \le n_L, \tag{31f}$$

$$\frac{dL_k^{(\mathrm{St},*)}}{dt} = \xi s_L L_k^{(\mathrm{St},-)}(t) + \delta L_{k-1}^{(\mathrm{St},*)}(t) - \delta L_k^{(\mathrm{St},*)}(t) - \alpha L_k^{(\mathrm{St},*)}(t) \qquad \text{for } 2 \le k \le n_L, \tag{31g}$$

$$\frac{dL_k^{(\mathrm{St},+)}}{dt} = \alpha L_k^{(\mathrm{St},*)}(t) + \delta L_{k-1}^{(\mathrm{St},+)}(t) - \delta L_k^{(\mathrm{St},+)}(t) \qquad \text{for } 2 \le k \le n_L, \tag{31h}$$

$$\frac{dL_1^{(\mathrm{Ri})}}{dt} = \gamma I_{n_I}^{(\mathrm{Ri})}(t) - \delta L_1^{(\mathrm{Ri})}(t), \tag{31i}$$

$$\frac{dL_k^{(\mathrm{Ri})}}{dt} = \delta L_{k-1}^{(\mathrm{Ri})}(t) - \delta L_k^{(\mathrm{Ri})}(t) \qquad \text{for } 2 \le k \le n_L. \tag{31j}$$

The dynamics of the recovered sub-populations are

$$\frac{dR^{(\mathrm{Ge})}}{dt} = \delta\big(1 - f_{\mathrm{Sick}} f_{\mathrm{Dead}}\big) L_{n_L}^{(\mathrm{Ge})}(t), \tag{32a}$$

$$\frac{dR^{(\mathrm{St,\text{-}})}}{dt} = \delta\big(1 - f_{\mathrm{Sick}} f_{\mathrm{Dead}}\big) L_{n_L}^{(\mathrm{St},-)}(t), \tag{32b}$$

$$\frac{dR^{(\mathrm{St,*})}}{dt} = \delta\big(1 - f_{\mathrm{Sick}} f_{\mathrm{Dead}}\big) L_{n_L}^{(\mathrm{St},*)}(t), \tag{32c}$$

$$\frac{dR^{(\mathrm{St,+})}}{dt} = \delta\big(1 - f_{\mathrm{Sick}} f_{\mathrm{Dead}}\big) L_{n_L}^{(\mathrm{St},+)}(t), \tag{32d}$$

$$\frac{dR^{(\mathrm{Ri})}}{dt} = \delta\left(1 - {f_{\mathrm{Sick}}}^{(\mathrm{Ri})} f_{\mathrm{Dead}}^{(\mathrm{Ri})}\right) L_{n_L}^{(\mathrm{Ri})}(t). \tag{32e}$$

Finally, the dynamics of dead individuals are given by

$$\frac{dD^{(\mathrm{Ge})}}{dt} = \delta f_{\mathrm{Sick}} f_{\mathrm{Dead}} L_{n_L}^{(\mathrm{Ge})}(t), \tag{33a}$$

$$\frac{dD^{(\mathrm{St,\text{-}})}}{dt} = \delta f_{\mathrm{Sick}} f_{\mathrm{Dead}} L_{n_L}^{(\mathrm{St},-)}(t), \tag{33b}$$

$$\frac{dD^{(\mathrm{St,*})}}{dt} = \delta f_{\mathrm{Sick}} f_{\mathrm{Dead}} L_{n_L}^{(\mathrm{St},*)}(t), \tag{33c}$$

$$\frac{dD^{(\mathrm{St,+})}}{dt} = \delta f_{\mathrm{Sick}} f_{\mathrm{Dead}} L_{n_L}^{(\mathrm{St},+)}(t), \tag{33d}$$

$$\frac{dD^{(\mathrm{Ri})}}{dt} = \delta {f_{\mathrm{Sick}}}^{(\mathrm{Ri})} f_{\mathrm{Dead}}^{(\mathrm{Ri})} L_{n_L}^{(\mathrm{Ri})}(t). \tag{33e}$$

#### Mathematical description of the demographic mixing matrix

The average numbers of daily contacts of an individual in the general sub-populations, among the LTCF employees, and the risk group are $n^{(\mathrm{Ge})}$, $n^{(\mathrm{St})}$, $n^{(\mathrm{Ri})}$, respectively. The total number of contacts in the three sub-populations are respectively the products $n^{(\mathrm{Ge})}N^{(\mathrm{Ge})}$, $n^{(\mathrm{St})}N^{(\mathrm{St})}$, and $n^{(\mathrm{Ri})}N^{(\mathrm{Ri})}$. For the general sub-population, $x$ and $y$ are the (conditional) probabilities that a contact occurs with and individual of the general sub-population or the sub-population of LTCF employees, respectively. Similarly for the LTCF staff, $u$ and $v$ are the probabilities that a contact occurs with the general-sub-population or the LTCF employees. Finally, for the risk group $p$ and $q$ are the probabilities of a contact with the general sub-population or the LTCF staff. Therefore, the matrix of all contacts becomes

$$M_{\mathrm{Tot}} := \begin{pmatrix} xn^{(\mathrm{Ge})}N^{(\mathrm{Ge})} & yn^{(\mathrm{Ge})}N^{(\mathrm{Ge})} & (1-x-y)n^{(\mathrm{Ge})}N^{(\mathrm{Ge})} \\ un^{(\mathrm{St})}N^{(\mathrm{St})} & vn^{(\mathrm{St})}N^{(\mathrm{St})} & (1-u-v)n^{(\mathrm{St})}N^{(\mathrm{St})} \\ pn^{(\mathrm{Ri})}N^{(\mathrm{Ri})} & qn^{(\mathrm{Ri})}N^{(\mathrm{Ri})} & (1-p-q)n^{(\mathrm{Ri})}N^{(\mathrm{Ri})} \end{pmatrix}. \tag{34}$$

The bidirectional nature of contacts implies that this matrix is symmetric and hence can be rewritten as

$$M_{\mathrm{Tot}} = \begin{pmatrix} xn^{(\mathrm{Ge})}N^{(\mathrm{Ge})} & yn^{(\mathrm{Ge})}N^{(\mathrm{Ge})} & (1-x-y)n^{(\mathrm{Ge})}N^{(\mathrm{Ge})} \\ yn^{(\mathrm{Ge})}N^{(\mathrm{Ge})} & vn^{(\mathrm{St})}N^{(\mathrm{St})} & (1-u-v)n^{(\mathrm{St})}N^{(\mathrm{St})} \\ (1-x-y)n^{(\mathrm{Ge})}N^{(\mathrm{Ge})} & (1-u-v)n^{(\mathrm{St})}N^{(\mathrm{St})} & (1-p-q)n^{(\mathrm{Ri})}N^{(\mathrm{Ri})} \end{pmatrix}. \tag{35}$$

This implies the relations

$$u = y\frac{n^{(\mathrm{Ge})}N^{(\mathrm{Ge})}}{n^{(\mathrm{St})}N^{(\mathrm{St})}}, \tag{36a}$$

$$p = (1-x-y)\frac{n^{(\mathrm{Ge})}N^{(\mathrm{Ge})}}{n^{(\mathrm{Ri})}N^{(\mathrm{Ri})}}, \tag{36b}$$

$$q = (1-u-v)\frac{n^{(\mathrm{St})}N^{(\mathrm{St})}}{n^{(\mathrm{Ri})}N^{(\mathrm{Ri})}}. \tag{36c}$$

The symmetry of the matrix constrains the choices of $n^{(\mathrm{Ge})}$, $n^{(\mathrm{St})}$, $n^{(\mathrm{Ri})}$, $x$, $y$, $u$, $v$, $p$, and $q$.

If individuals in the whole population would encounter randomly, which is not the case due to the specific contact behaviour, the probabilities of encounters between the sub-populations was

$$R := \begin{pmatrix} \left(\frac{N^{(\mathrm{Ge})}}{N}\right)^2 & \frac{N^{(\mathrm{Ge})}N^{(\mathrm{St})}}{N^2} & \frac{N^{(\mathrm{Ge})}N^{(\mathrm{Ri})}}{N^2} \\ \frac{N^{(\mathrm{Ge})}N^{(\mathrm{St})}}{N^2} & \left(\frac{N^{(\mathrm{St})}}{N}\right)^2 & \frac{N^{(\mathrm{St})}N^{(\mathrm{Ri})}}{N^2} \\ \frac{N^{(\mathrm{Ge})}N^{(\mathrm{Ri})}}{N^2} & \frac{N^{(\mathrm{St})}N^{(\mathrm{Ri})}}{N^2} & \left(\frac{N^{(\mathrm{Ri})}}{N}\right)^2 \end{pmatrix}. \tag{37}$$

The mixing matrix $X$ corrects for the dependency in the behavior of encounters such that the matrix of all contacts emerges as the scalar product (element-wise product) of the matrix $R$ and the mixing matrix $X$, i.e., $M_{\mathrm{Tot}} = R \odot X$. Thus the mixing matrix $X$ is given by

$$X := M_{\mathrm{Tot}} \odot (1/R) = \begin{pmatrix} xn^{(\mathrm{Ge})}\frac{N^2}{N^{(\mathrm{Ge})}} & yn^{(\mathrm{Ge})}\frac{N^2}{N^{(\mathrm{St})}} & (1-x-y)n^{(\mathrm{Ge})}\frac{N^2}{N^{(\mathrm{Ri})}} \\ un^{(\mathrm{St})}\frac{N^2}{N^{(\mathrm{Ge})}} & vn^{(\mathrm{St})}\frac{N^2}{N^{(\mathrm{St})}} & (1-u-v)n^{(\mathrm{St})}\frac{N^2}{N^{(\mathrm{Ri})}} \\ pn^{(\mathrm{Ri})}\frac{N^2}{N^{(\mathrm{Ge})}} & qn^{(\mathrm{Ri})}\frac{N^2}{N^{(\mathrm{St})}} & (1-p-q)n^{(\mathrm{Ri})}\frac{N^2}{N^{(\mathrm{Ri})}} \end{pmatrix}, \tag{38}$$

where $1/R$ denotes the matrix which has the reciprocal entries of $R$.

##### General contact reduction and restricted facility access

General contact reduction is sustained in a time-dependent fashion. At each time point, the amount of contacts being reduced depends on the characteristics of the interactions between sub-populations. This changes the matrix $M_{\mathrm{Tot}}$ to $\tilde{M}_{\mathrm{Tot}}(t)$. Let $p_{\mathrm{Cont}}^{(i,j)}(t) = p_{\mathrm{Cont}}^{(j,i)}(t)$ be the fraction of contacts that is reduced between sub-populations $i$ and $j$ at time $t$ ($i, j =$ (Ge), (St), (Ri)). The actual numbers of these reductions depend on the type of interventions being sustained. The matrix $\tilde{M}_{\mathrm{Tot}}(t)$ is given by

$$\tilde{M}_{\mathrm{Tot}}(t) = M_{\mathrm{Tot}} \odot \begin{pmatrix} 1-p_{\mathrm{Cont}}^{(\mathrm{Ge,Ge})}(t) & 1-p_{\mathrm{Cont}}^{(\mathrm{Ge,St})}(t) & 1-p_{\mathrm{Cont}}^{(\mathrm{Ge,Ri})}(t) \\ 1-p_{\mathrm{Cont}}^{(\mathrm{Ge,Ri})}(t) & 1-p_{\mathrm{Cont}}^{(\mathrm{St,St})}(t) & 1-p_{\mathrm{Cont}}^{(\mathrm{St,Ri})}(t) \\ 1-p_{\mathrm{Cont}}^{(\mathrm{Ge,Ri})}(t) & 1-p_{\mathrm{Cont}}^{(\mathrm{Ri,St})}(t) & 1-p_{\mathrm{Cont}}^{(\mathrm{Ri,Ri})}(t) \end{pmatrix}. \tag{39}$$

The mixing matrix $X$ becomes a function of time defined by

$$X(t) = \tilde{M}_{\mathrm{Tot}}(t) \odot (1/R). \tag{40}$$

#### The basic reproduction number and the next generation matrix

The normal classical definition of the basic reproductive number $R_0$ is the average number of infections caused by an infected individual in a completely susceptible population. This definition does not make sense in a heterogeneous subdivided population, in this case it is rather the average number of infections caused by an average infected individual [?] in a susceptible population without any disease-control interventions. In this case $R_0$ is derived as the maximum eigenvalue of the next-generation matrix (NGM).

The NGM is derived by first linearizing the reduced system of ODEs assuming no control interventions and all individuals to be susceptible. In particular, the reduced system is obtained by retaining only those differential equations from the original system that describe the states of infected individuals, which are relevant in the absence of interventions. Let $\boldsymbol{x}$ be the vector of all states in the reduced system. The Jacobian of this system is derived at the state, in which all individuals are susceptible, denoted $\boldsymbol{x}_0$. The Jacobian is split into one matrix describing transmission and one matrix describing transitions between infected states. Equivalently, vector-valued functions $F(\boldsymbol{x})$ and $V(\boldsymbol{x})$, describing transmission and transitions in the reduced system can be calculated, whose sum of Jacobian matrices, $\dfrac{\partial F}{\partial \boldsymbol{x}}$ and $\dfrac{\partial V}{\partial \boldsymbol{x}}$, equals the Jacobian of the reduced system. The NGM is calculated as

$$G := -\left[\frac{\partial F}{\partial \boldsymbol{x}}(\boldsymbol{x}_0)\right]\left[\frac{\partial V}{\partial \boldsymbol{x}}(\boldsymbol{x}_0)\right]^{-1}. \tag{41}$$

Finally, $R_0$ is obtained as the spectral radius of the matrix $G$, i.e., as the maximum absolute eigenvalue (cf. [?]). In mathematical terms

$$R_0 := \rho(G) := \max_i |\lambda_i(G)|, \tag{42}$$

where $\lambda_i(G)$ denote the eigenvalues of $G$. Note that $\rho(G)$ is a function of $\bar{R}_0^{(\mathrm{adj})}$. It has to be chosen such that

$$\bar{R}_0 = \frac{\max_i |\lambda_i(G)|}{1 + a\cos\left(-2\pi\frac{t_{R_{0_{\max}}}}{365}\right)}$$

holds.

In our case, the reduced system omits equations (27), (28e), (28f), (28h), (28i), (29d), (29e), (29g) (29h), (30d), (30e), (30g) (30h), (31d), (31e), (31g) (31h), (32), and (33).

The function $F$ becomes

$$F(\boldsymbol{x}) = \begin{pmatrix} \left(\lambda_{\text{Ge}}(t) + \lambda_{\text{Ext}}\right) \dfrac{S^{(\text{Ge})}(t)}{N} \\ 0 \\ \vdots \\ 0 \\ \left(\lambda_{\text{St}}(t) + \lambda_{\text{Ext}}\right) \dfrac{S^{(\text{St})}(t)}{N} \\ 0 \\ \vdots \\ 0 \\ \lambda_{\text{Ri}}(t) \dfrac{S^{(\text{Ri})}(t)}{N} \\ 0 \\ \vdots \\ 0 \end{pmatrix} = \begin{pmatrix} F_{E_1^{(\text{Ge})}} \\ 0 \\ \vdots \\ 0 \\ F_{E_1{}^{(\text{St},-)}} \\ 0 \\ \vdots \\ 0 \\ F_{E_1^{(\text{Ri})}} \\ 0 \\ \vdots \\ 0 \end{pmatrix}. \tag{43}$$

The non-vanishing entries of the Jacobian of $F$ are

$$\frac{\partial F_{E_1^{(l)}}}{\partial P_k^{(j)}} := \beta_P(t) x_{i,j} \frac{S^{(i)}}{N} \qquad \text{for} \quad k = 1, \dots, n_P, \tag{44a}$$

$$\frac{\partial F_{E_1^{(l)}}}{\partial I_k^{(j)}} := \beta_I(t) x_{i,j} \frac{S^{(i)}}{N} \qquad \text{for} \quad k = 1, \dots, n_I, \tag{44b}$$

$$\frac{\partial F_{E_1^{(l)}}}{\partial L_k^{(j)}} := \beta_L(t) x_{i,j} \frac{S^{(i)}}{N} \qquad \text{for} \quad k = 1, \dots, n_L, \tag{44c}$$

for $i$, $j$ = ‘Ge’, ‘St’, ‘Ri’ and $l = i$ except for $i$ = ‘St’, in which case $l$ = ‘St,-’.

Furthermore,

$$\big(V_1(\boldsymbol{x}),\ldots,V_{3n_E}(\boldsymbol{x})\big)=\begin{pmatrix} -\varepsilon E_1^{(\mathrm{Ge})}(t)\\ \vdots\\ \varepsilon E_{k-1}^{(\mathrm{Ge})}(t)-\varepsilon E_k^{(\mathrm{Ge})}(t)\\ \vdots\\ -\varepsilon E_1^{(\mathrm{St},-)}(t)\\ \vdots\\ \varepsilon E_{k-1}^{(\mathrm{St},-)}(t)-\varepsilon E_k^{(\mathrm{St},-)}(t)\\ \vdots\\ -\varepsilon E_1^{(\mathrm{Ri})}(t)\\ \vdots\\ \varepsilon E_{k-1}^{(\mathrm{Ri})}(t)-\varepsilon E_k^{(\mathrm{Ri})}(t)\\ \vdots \end{pmatrix}, \tag{45a}$$

$$\big(V_{3n_E+1}(\boldsymbol{x}),\ldots,V_{3(n_E+n_P)}(\boldsymbol{x})\big)=\begin{pmatrix} \varepsilon E_{n_E}^{(\mathrm{Ge})}(t)-\varphi P_1^{(\mathrm{Ge})}(t)\\ \vdots\\ \varphi P_{k-1}^{(\mathrm{Ge})}(t)-\varphi P_k^{(\mathrm{Ge})}(t)\\ \vdots\\ \varepsilon E_{n_E}^{(\mathrm{St},-)}(t)-\varphi P_1^{(\mathrm{St},-)}(t)\\ \vdots\\ \varphi P_{k-1}^{(\mathrm{St},-)}(t)-\varphi P_k^{(\mathrm{St},-)}(t)\\ \vdots\\ \varepsilon E_{n_E}^{(\mathrm{Ri})}(t)-\varphi P_1^{(\mathrm{Ri})}(t)\\ \vdots\\ \varphi P_{k-1}^{(\mathrm{Ri})}(t)-\varphi P_k^{(\mathrm{Ri})}(t)\\ \vdots \end{pmatrix}, \tag{45b}$$

$$\big(V_{3(n_E+n_P)+1}(\boldsymbol{x}),\dots,V_{3(n_E+n_P+n_I)}(\boldsymbol{x})\big)=\begin{pmatrix}\varphi P_{n_P}^{(\mathrm{Ge})}(t)-\gamma I_1^{(\mathrm{Ge})}(t)\\ \vdots\\ \gamma I_{k-1}^{(\mathrm{Ge})}(t)-\gamma I_k^{(\mathrm{Ge})}(t)\\ \vdots\\ \varphi P_{n_P}^{(\mathrm{St},-)}(t)-\gamma I_1^{(\mathrm{St},-)}(t)\\ \vdots\\ \gamma I_{k-1}^{(\mathrm{St},-)}(t)-\gamma I_k^{(\mathrm{St},-)}(t)\\ \vdots\\ \varphi P_{n_P}^{(\mathrm{Ri})}(t)-\gamma I_1^{(\mathrm{Ri})}(t)\\ \vdots\\ \gamma I_{k-1}^{(\mathrm{Ri})}(t)-\gamma I_k^{(\mathrm{Ri})}(t)\\ \vdots\end{pmatrix}, \tag{45c}$$

$$\big(V_{3(n_E+n_P+n_I)+1}(\boldsymbol{x}),\dots,V_{3(n_E+n_P+n_I+n_L)}(\boldsymbol{x})\big)=\begin{pmatrix}\gamma I_{n_I}^{(\mathrm{Ge})}(t)-\delta L_1^{(\mathrm{Ge})}(t)\\ \vdots\\ \delta L_{k-1}^{(\mathrm{Ge})}(t)-\delta L_k^{(\mathrm{Ge})}(t)\\ \vdots\\ \gamma I_{n_I}^{(\mathrm{St},-)}(t)-\delta L_1^{(\mathrm{St},-)}(t)\\ \vdots\\ \delta L_{k-1}^{(\mathrm{St},-)}(t)-\delta L_k^{(\mathrm{St},-)}(t)\\ \vdots\\ \gamma I_{n_I}^{(\mathrm{Ri})}(t)-\delta L_1^{(\mathrm{Ri})}(t)\\ \vdots\\ \delta L_{k-1}^{(\mathrm{Ri})}(t)-\delta L_k^{(\mathrm{Ri})}(t)\\ \vdots\end{pmatrix}. \tag{45d}$$

The derivatives of the $V(\boldsymbol{x})$ are straightforward and are omitted here. Deriving (41) and (42) from (44) and the derivatives of (45) is straightforward.

To find the proper values of $\bar{R}_0^{(\mathrm{adj})}$, let $\tilde{G}$ denote the matrix

$$\tilde{G}:=\frac{1}{\bar{R}_0^{(\mathrm{adj})}\left(1+a\cos\left(-2\pi\frac{t_{R_{0_{\max}}}}{365}\right)\right)}G, \tag{46}$$

which is independent of $\bar{R}_0^{(\mathrm{adj})}$. Then

$$\bar{R}_0^{(\mathrm{adj})}=\frac{\bar{R}_0}{\max_i|\lambda_i(\tilde{G})|}. \tag{47}$$
