## Supplementary material for "Preventing COVID-19 spread in closed facilities by regular testing of employees – an efficient intervention in long-term care facilities and prisons?": Table S1

**S1 Table.** (Sub-) population sizes of Germany (GER) and the USA chosen in simulations.

| Parameter | Description | GER | USA |
|---|---|---|---|
| $N$ | Total population size | 83,000,000 | 331,000,000 |
| $N^{(\mathrm{Ge})}$ | Size of general sub-population | 81,800,000 | 329,177,000 |
| $N^{(\mathrm{St})}$ | Number of LTCF employees | 500,000 | 423,000 |
| $N^{(\mathrm{Ri})}$ | Size of risk group | 700,000 | 1,400,000 |
