## Supplementary material for "Preventing COVID-19 spread in closed facilities by regular testing of employees – an efficient intervention in long-term care facilities and prisons?": Fig S2

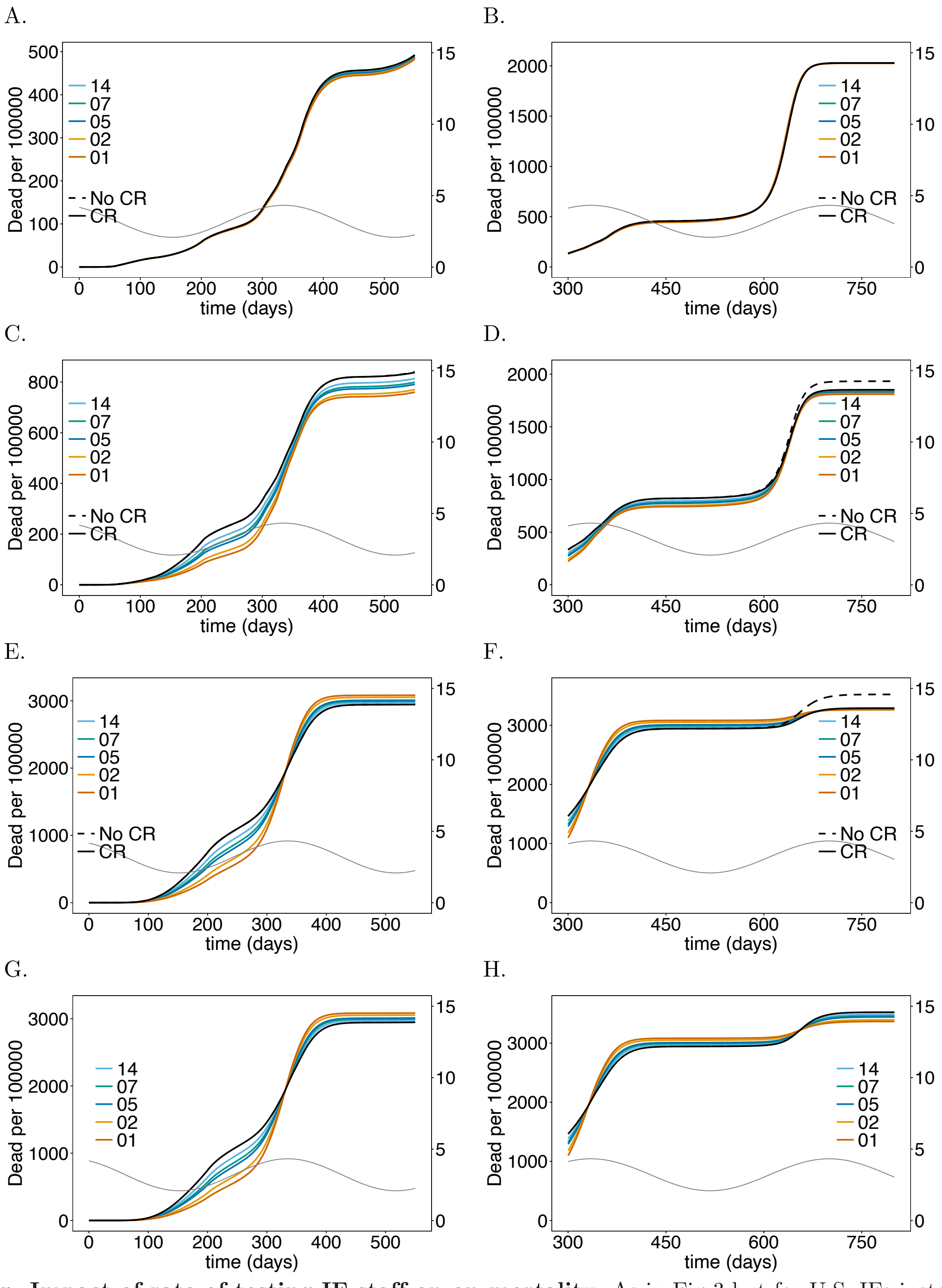


**S2 Fig. Impact of rate of testing IF staff on on mortality**: As in Fig 3 but for U.S. IFs instead of German LTCFs. Parameters for contact reduction are given in S7 Table.
