## Supplementary material for "Preventing COVID-19 spread in closed facilities by regular testing of employees – an efficient intervention in long-term care facilities and prisons?": Table S2

**S2 Table.** Summary of model parameters and their choices for numerical simulations for Germany (GER) and USA.

| Parameter | Description | Value | |
|---|---|---|---|
| $n_E$ | Number of latent phase Erlang states | 16 | |
| $n_P$ | Number of prodromal Erlang states | 16 | |
| $n_I$ | Number of fully infectious Erlang states | 16 | |
| $n_L$ | Number of late infectious Erlang states | 16 | |
| $D_E$ | Average duration of latency period | 3.7 days | |
| $D_P$ | Average duration of prodromal period | 1 day | |
| $D_I$ | Average duration of fully infectious period | 5 days | |
| $D_L$ | Average duration of late infectious period | 5 days | |
| $\varepsilon$ | Transition rate of latent states | $n_E/D_E$ | |
| $\varphi$ | Transition rate of prodromal states | $n_P/D_P$ | |
| $\gamma$ | Transition rate of fully infectious states | $n_I/D_I$ | |
| $\delta$ | Transition rate of late infectious states | $n_L/D_L$ | |
| $1/\alpha$ | Waiting time for test results in days | 1/2, 1, 2, 3, 4 | |
| $1/\xi$ | Average frequency of testing in days | 1, 2, 5, 7, 14 | |
| | | GER | USA |
| $f_{\text{Sick}}$ | Fraction of symptomatic (sick) infections in Ge and St | 58% | 58% |
| $f_{\text{Sick}}^{(\text{Ri})}$ | Fraction of symptomatic (sick) infections in Ri | 60% | 43% |
| $f_{\text{Dead}}$ | Fraction of sick ind. in Ge and St, who die from the disease | 1.6% | 4% |
| $f_{\text{Dead}}^{(\text{Ri})}$ | Fraction of sick ind. in Ri, who die from the disease | 20% | 11% |
| $f_{\text{Iso}}$ | Fraction of sick ind. who go to isolation | 58% | 48% |

General sub-population (Ge), LTCF employees (St), risk group (Ri).
