## Supplementary material for "Preventing COVID-19 spread in closed facilities by regular testing of employees – an efficient intervention in long-term care facilities and prisons?": Table S3

**S3 Table.** Summary of variables describing sub-population sizes in non-infectious (sub-)states in Germany (GER) and the USA.

| Name | Description | Initial value | |
|---|---|---|---|
| | | GER | USA |
| $S^{(\mathrm{Ge})}$ | No. susceptibles in Ge | 81,799,800 | 329,176,925 |
| $S^{(\mathrm{St})}$ | No. susceptibles in St | 500,000 | 423,000 |
| $S^{(\mathrm{Ri})}$ | No. susceptibles in Ri | 700,000 | 1,400,000 |
| $E_k^{(\mathrm{Ge})}$ | No. infected Ge in $k$th latent state ($1 \leq k \leq n_E$) | | 0 |
| $E_k^{(\mathrm{St},-)}$ | No. infected undetected St in $k$th latent state ($1 \leq k \leq n_E$) | | 0 |
| $E_k^{(\mathrm{St},*)}$ | No. St in $k$th latent state, whose test results will be pos. ($1 \leq k \leq n_E$) | | 0 |
| $E_k^{(\mathrm{St},+)}$ | No. pos. tested St in $k$th latent state ($1 \leq k \leq n_E$) | | 0 |
| $E_k^{(\mathrm{Ri})}$ | No. Ri in $k$th latent state ($1 \leq k \leq n_E$) | | 0 |
| $E_{\mathrm{Sum}}^{(\mathrm{Ge})}$ | Total No. of Ge in latent states | | 0 |
| $E_{\mathrm{Sum}}^{(\mathrm{St},-)}$ | Total No. undetected infected St in latent states | | 0 |
| $E_{\mathrm{Sum}}^{(\mathrm{St},*)}$ | Total No. of St in latent states, whose test results will be pos. | | 0 |
| $E_{\mathrm{Sum}}^{(\mathrm{St},+)}$ | Total No.of pos. tested St in latent states | | 0 |
| $E_{\mathrm{Sum}}^{(\mathrm{Ri})}$ | Total No. of Ri in latent states | | 0 |
| $R^{(\mathrm{Ge})}$ | No. recovered in Ge | | 0 |
| $R^{(\mathrm{St},-)}$ | No. recovered in St, whose infections were undetected | | 0 |
| $R^{(\mathrm{St},*)}$ | No. St that recovered, before pos. test result returned | | 0 |
| $R^{(\mathrm{St},+)}$ | No. recovered St that were pos. tested | | 0 |
| $R^{(\mathrm{Ri})}$ | No. recovered in Ri | | 0 |
| $D^{(\mathrm{Ge})}$ | No. dead in Ge | | 0 |
| $D^{(\mathrm{St},-)}$ | No. dead in St, whose infections were undetected | | 0 |
| $D^{(\mathrm{St},*)}$ | No. St that died, before pos. test result returned | | 0 |
| $D^{(\mathrm{St},+)}$ | No. dead St that were pos. tested | | 0 |
| $D^{(\mathrm{Ri})}$ | No. dead in Ri | | 0 |

Description of variables and their initial values chosen for the simulations.
