## Supplementary material for "Preventing COVID-19 spread in closed facilities by regular testing of employees – an efficient intervention in long-term care facilities and prisons?": Fig S4

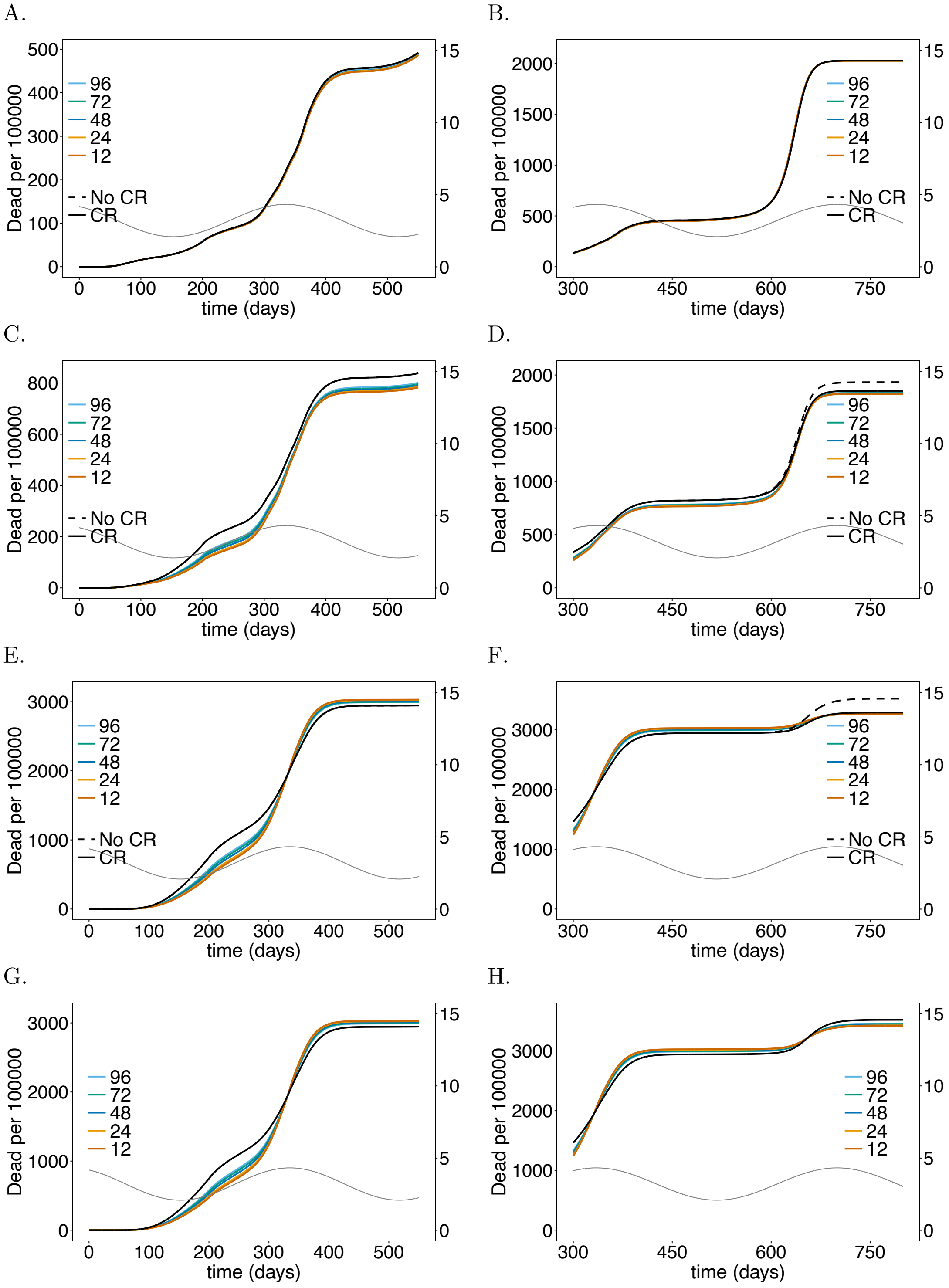


**S4 Fig. Impact of processing time of testing IF staff on on mortality:** As in Fig 5 but for U.S. IFs instead of German LTCFs. Parameters for contact reduction are given in S7 Table.
