## Supplementary material for "Preventing COVID-19 spread in closed facilities by regular testing of employees – an efficient intervention in long-term care facilities and prisons?": Table S4

**S4 Table.** Summary of variables describing sub-population sizes in infectious (sub-)states in Germany (GER) and the USA.

| Name | Description | Initial value | |
|---|---|---|---|
| $P_k^{(\mathrm{Ge})}$ | No. infected Ge in $k$th prodromal state ($1 \le k \le n_P$) | | 0 |
| $P_k^{(\mathrm{St},-)}$ | No. infected undetected St in $k$th prodromal state ($1 \le k \le n_P$) | | 0 |
| $P_k^{(\mathrm{St},*)}$ | No. St in $k$th prodromal state, whose test results will be pos. ($1 \le k \le n_P$) | | 0 |
| $P_k^{(\mathrm{St},+)}$ | No. pos. tested St in $k$th prodromal state ($1 \le k \le n_P$) | | 0 |
| $P_k^{(\mathrm{Ri})}$ | No. Ri in $k$th prodromal state ($1 \le k \le n_P$) | | 0 |
| $P_{\mathrm{Sum}}^{(\mathrm{Ge})}$ | Total No. of Ge in prodromal states | | 0 |
| $P_{\mathrm{Sum}}^{(\mathrm{St},-)}$ | Total No. undetected infected St in prodromal states | | 0 |
| $P_{\mathrm{Sum}}^{(\mathrm{St},*)}$ | Total No. of St in prodromal states, whose test results will be pos. | | 0 |
| $P_{\mathrm{Sum}}^{(\mathrm{St},+)}$ | Total No. of pos. tested St in prodromal states | | 0 |
| $P_{\mathrm{Sum}}^{(\mathrm{Ri})}$ | Total No. of Ri in prodromal states | | 0 |
| | | GER | USA |
| $I_1^{(\mathrm{Ge})}$ | No. infected Ge in 1st fully infectious state | 200 | 75 |
| $I_k^{(\mathrm{Ge})}$ | No. infected Ge in $k$th fully infectious state ($2 \le k \le n_I$) | 0 | 0 |
| $I_{\mathrm{Sum}}^{(\mathrm{Ge})}$ | Total No. of Ge in fully infectious states | 200 | 75 |
| $I_k^{(\mathrm{St},-)}$ | No. infected undetected St in $k$th fully infectious state ($1 \le k \le n_I$) | | 0 |
| $I_k^{(\mathrm{St},*)}$ | No. St in $k$th fully infectious state, whose test results will be pos. ($1 \le k \le n_I$) | | 0 |
| $I_k^{(\mathrm{St},+)}$ | No. pos. tested St in $k$th fully infectious state ($1 \le k \le n_I$) | | 0 |
| $I_k^{(\mathrm{Ri})}$ | No. Ri in $k$th fully infectious state ($1 \le k \le n_I$) | | 0 |
| $I_{\mathrm{Sum}}^{(\mathrm{St},-)}$ | Total No. undetected infected St in fully infectious states | | 0 |
| $I_{\mathrm{Sum}}^{(\mathrm{St},*)}$ | Total No. of St in fully infectious states, whose test results will be pos. | | 0 |
| $I_{\mathrm{Sum}}^{(\mathrm{St},+)}$ | Total No.of pos. tested St in fully infectious states | | 0 |
| $I_{\mathrm{Sum}}^{(\mathrm{Ri})}$ | Total No. of Ri in fully infectious states | | 0 |
| $L_k^{(\mathrm{Ge})}$ | No. infected Ge in $k$th late infectious state ($1 \le k \le n_L$) | | 0 |
| $L_k^{(\mathrm{St},-)}$ | No. infected undetected St in $k$th late infectious state ($1 \le k \le n_L$) | | 0 |
| $L_k^{(\mathrm{St},*)}$ | No. St in $k$th late infected state, whose test results will be pos. ($1 \le k \le n_L$) | | 0 |
| $L_k^{(\mathrm{St},+)}$ | No. pos. tested St in $k$th late infectious state ($1 \le k \le n_L$) | | 0 |
| $L_k^{(\mathrm{Ri})}$ | No. Ri in $k$th late infectious state ($1 \le k \le n_L$) | | 0 |
| $L_{\mathrm{Sum}}^{(\mathrm{Ge})}$ | Total No. of Ge in late infectious states | | 0 |
| $L_{\mathrm{Sum}}^{(\mathrm{St},-)}$ | Total No. infected undetected individuals in late infectious state in St | | 0 |
| $L_{\mathrm{Sum}}^{(\mathrm{St},*)}$ | Total No. of St in late infected states, whose test results will be pos. | | 0 |
| $L_{\mathrm{Sum}}^{(\mathrm{St},+)}$ | Total No.of pos. tested St in late infectious states | | 0 |
| $L_{\mathrm{Sum}}^{(\mathrm{Ri})}$ | Total No. of Ri in late infectious states | | 0 |

Description of variables and their initial values chosen for the simulations.
