## Supplementary material for "Preventing COVID-19 spread in closed facilities by regular testing of employees – an efficient intervention in long-term care facilities and prisons?": Fig S5

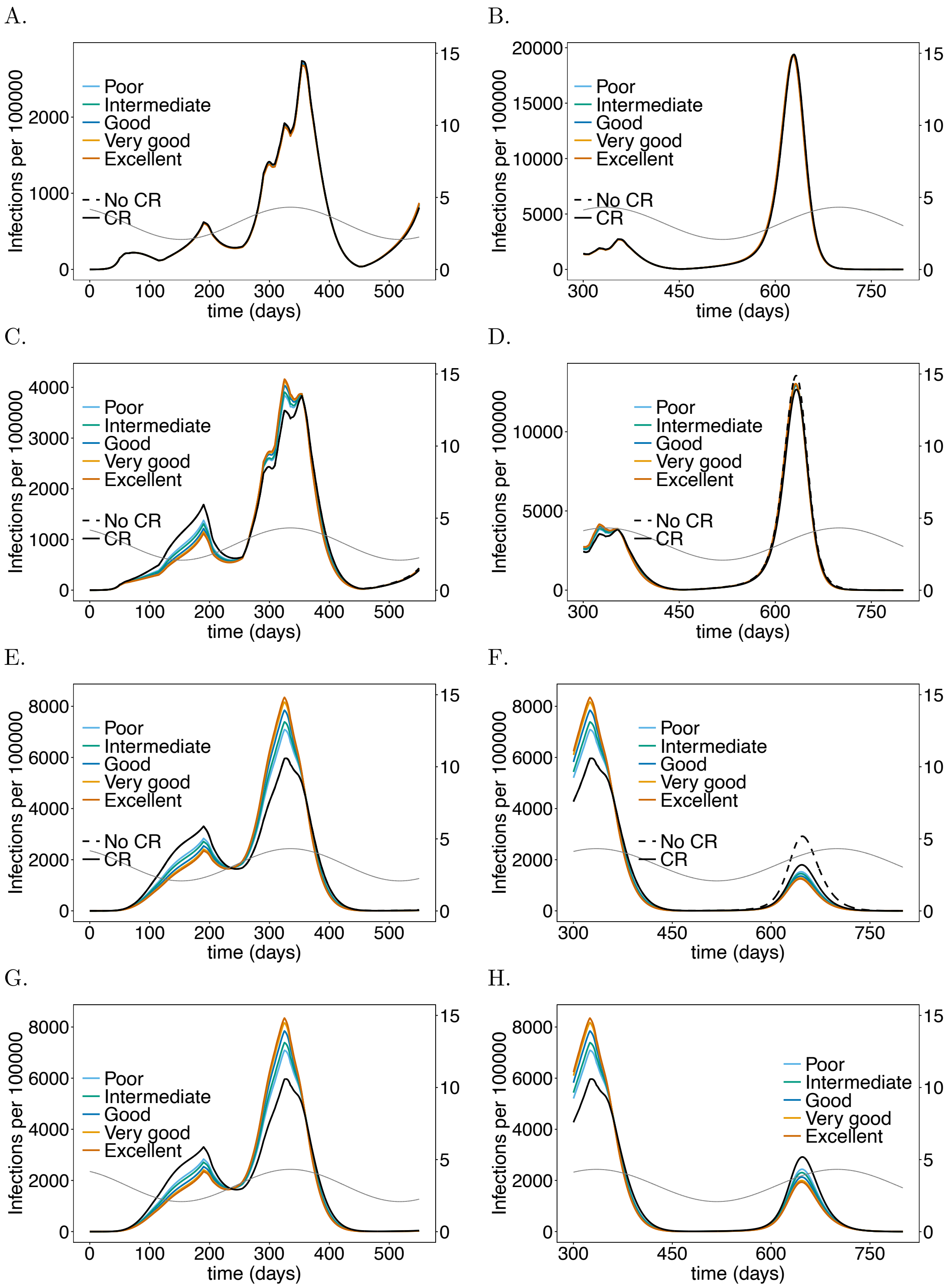


**S5 Fig. Impact of sensitivity when testing IF staff on the number of infections**: As in Fig 6 but for U.S. IFs instead of German LTCFs. Parameters for contact reduction are given in S7 Table.
