## Supplementary material for "Preventing COVID-19 spread in closed facilities by regular testing of employees – an efficient intervention in long-term care facilities and prisons?": Table S5

**S5 Table.** Summary of model parameters describing the contact behavior for Germany (GER) and USA.

| Name | Description | Value/Eq. | |
|---|---|---|---|
| | | GER | USA |
| $\lambda_{\text{Ext}}$ | Infections from outside of the population | 45/day | 50/day |
| $\bar{R}_0$ | Annual average basic reproduction number | 3.4 | 3.2 |
| $a$ | Amplitude of the seasonal fluctuation of $R_0$ | 0.43 | 0.35 |
| $t_{R_{0\max}}$ | Day when $R_0$ reaches its maximum | 300 | 335 |
| $t_{\text{Iso}_1}$ | Day of beginning of case isolation measures | 30 | 20 |
| $t_{\text{Iso}_2}$ | Day of end of case isolation measures | 750 | 800 |
| $Q_{\max}$ | Maximum capacity of isolation units per 10 000 | 200 | 30 |
| $n^{(\text{Ge})}$ | Average number of daily contacts of an individual in Ge | 50 | 60 |
| $n^{(\text{St})}$ | Average number of daily contacts of an individual in St | 50 | 60 |
| $n^{(\text{Ri})}$ | Average number of daily contacts of an individual in Ri | 30 | 60 |
| $x$ | prob. for individual in Ge to meet with individual in Ge | 99.50% | 99.92% |
| $y$ | prob. for individual in Ge to meet with individual in St | 0.30% | 0.07% |
| $u$ | prob. for individual in St to meet with individual in Ge | 49.08% | 54.47% |
| $v$ | prob. for individual in the St to meet individual of the St | 20.0% | 20.0% |
| $p$ | prob. for individual in Ri to meet individual of the Ge | 38.95% | 2.35% |
| $q$ | prob. for individual in Ri to meet individual in St | 36.81% | 7.71% |
| $c_P$ | Relative infectiousness in prodromal period | | 0.5 |
| $c_I$ | Relative infectiousness in fully infectious stage | | 1 |
| $c_L$ | Relative infectiousness in late infectious stage | | 0.5 |
| $\beta_P(t)$ | Seasonally varying effective contact rate of prodromal ind. | | (23a) |
| $\beta_I(t)$ | Seasonally varying effective contact rate of fully infectious ind.s | | (23b) |
| $\beta_L(t)$ | Seasonally varying effective contact rate of late infectious ind. | | (23c) |
| $p_{\text{Home}}$ | Contact reduction in home isolation | | 75% |

Description of parameters and their initial values chosen for the simulations. Equation numbers refer to S1 Appendix
