## Supplementary material for "Preventing COVID-19 spread in closed facilities by regular testing of employees – an efficient intervention in long-term care facilities and prisons?": Fig S6

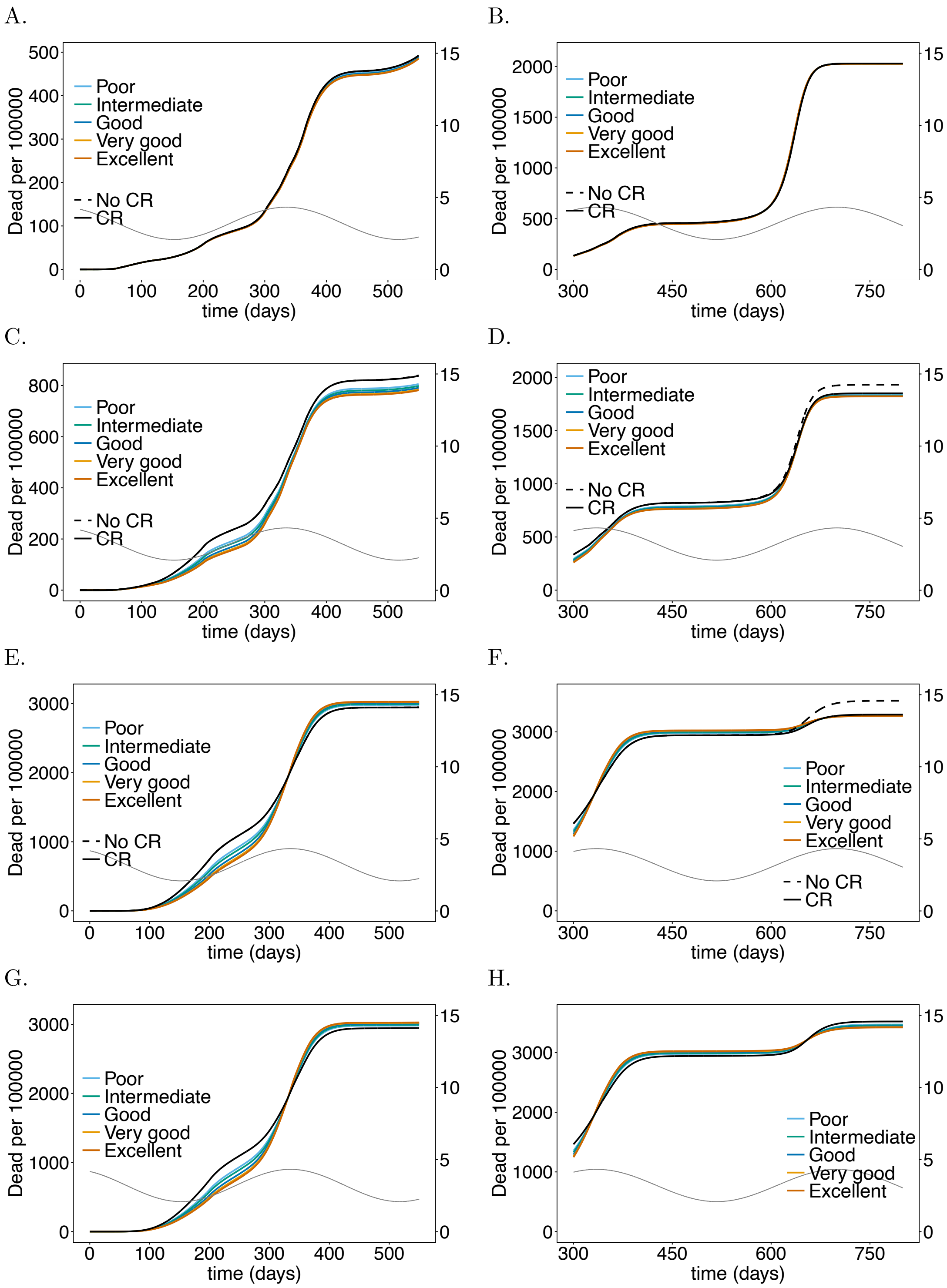


**S6 Fig. Impact of sensitivity when testing IF staff on mortality**: As in Fig 7 but for U.S. IFs instead of German LTCFs. Parameters for contact reduction are given in S7 Table.
