## Supplementary material for "Preventing COVID-19 spread in closed facilities by regular testing of employees – an efficient intervention in long-term care facilities and prisons?": Table S6

**S6 Table.** Contact reduction parameters chosen for the simulations of Germany.

| Parameter | Description | $I_1$ | $I_2$ | $I_3$ | $I_4$ | $I_5$ | $I_6$ |
|---|---|---|---|---|---|---|---|
| $t_{\text{Dist}_n}$-$t_{\text{Dist}_{n+1}}$ | Time intervals of gen. cont. red. | 40-82 | 82-246 | 246-280 | 280-380 | 380-450 | 450-750 |
| | Fraction of avoided contacts in time intervals $I_k$ between | | | | | | |
| $p_{\text{Cont}}^{(\text{Ge,Ge})}$ | Ge and Ge | 0.70 | 0.40 | 0.50 | 0.68 | 0.50 | 0 |
| $p_{\text{Cont}}^{(\text{Ge,St})}$ | Ge and St | 0.70 | 0.40 | 0.50 | 0.68 | 0.50 | 0 |
| $p_{\text{Cont}}^{(\text{Ge,Ri})}$ | Ge and Ri | 0.60 | 0.20 | 0.40 | 0.50 | 0.40 | 0 |
| $p_{\text{Cont}}^{(\text{St,St})}$ | St and St | 0.60 | 0.40 | 0.50 | 0.50 | 0.50 | 0.50 |
| $p_{\text{Cont}}^{(\text{St,Ri})}$ | St and Ri | 0.35 | 0.20 | 0.20 | 0.35 | 0.35 | 0.35 |
| $p_{\text{Cont}}^{(\text{Ri,Ri})}$ | Ri and Ri | 0.40 | 0.20 | 0.30 | 0.40 | 0.40 | 0.30 |
