## Supplementary material for "Preventing COVID-19 spread in closed facilities by regular testing of employees – an efficient intervention in long-term care facilities and prisons?": Table S7

**S7 Table.** Contact reduction parameters chosen for the simulations of USA.

| Parameter | $I_1$ | $I_2$ | $I_3$ | $I_4$ | $I_5$ | $I_6$ | $I_7$ | $I_8$ | $I_9$ | $I_{10}$ | $I_{11}$ |
|---|---|---|---|---|---|---|---|---|---|---|---|
| $t_{\mathrm{Dist}_n}$-$t_{\mathrm{Dist}_{n+1}}$ | 50-115 | 115-190 | 190-255 | 255-290 | 290-309 | 309-316 | 316-325 | 325-335 | 335-354 | 354-450 | 450-800 |
| | Fraction of avoided contacts in time intervals $I_k$ between | | | | | | | | | | |
| $p_{\mathrm{Cont}}^{(\mathrm{Ge,Ge})}$ | 0.55 | 0.22 | 0.55 | 0.45 | 0.65 | 0.55 | 0.60 | 0.70 | 0.55 | 0.70 | 0 |
| $p_{\mathrm{Cont}}^{(\mathrm{Ge,St})}$ | 0.55 | 0.22 | 0.55 | 0.45 | 0.65 | 0.55 | 0.60 | 0.70 | 0.55 | 0.70 | 0 |
| $p_{\mathrm{Cont}}^{(\mathrm{Ge,Ri})}$ | 0.55 | 0.22 | 0.55 | 0.45 | 0.65 | 0.55 | 0.60 | 0.70 | 0.55 | 0.70 | 0 |
| $p_{\mathrm{Cont}}^{(\mathrm{St,St})}$ | 0.37 | 0.05 | 0.50 | 0.30 | 0.55 | 0.40 | 0.50 | 0.55 | 0.45 | 0.55 | 0.50 |
| $p_{\mathrm{Cont}}^{(\mathrm{St,Ri})}$ | 0.20 | 0.05 | 0.50 | 0.25 | 0.45 | 0.30 | 0.35 | 0.45 | 0.35 | 0.45 | 0.40 |
| $p_{\mathrm{Cont}}^{(\mathrm{Ri,Ri})}$ | 0.05 | 0.00 | 0.25 | 0.20 | 0.25 | 0.22 | 0.20 | 0.30 | 0.20 | 0.25 | 0.20 |
